# Dynamic Substates of the Default Mode Network Are Associated with Mystical Experiences and Clinical Outcomes after Magnesium-Ibogaine Treatment in Veterans with Traumatic Brain Injury

**DOI:** 10.64898/2026.07.20.26358512

**Authors:** Kenneth Shinozuka, Clayton Olash, Tracy Han, Azeezat Azeez, Malvika Sridhar, Andrew D. Geoly, Catherine Daye, Saron Hunegnaw, Kirsten N. Cherian, Jackob N. Keynan, Randi E. Brown, Derrick M. Buchanan, John P. Coetzee, Ian H. Kratter, Raag D. Airan, Martijn Arns, Maheen M. Adamson, Manish Saggar, Cammie Rolle

## Abstract

Preliminary evidence suggests that the combination of magnesium and ibogaine, an atypical psychedelic, may be a promising treatment for post-traumatic stress disorder (PTSD), opioid use disorder, and traumatic brain injury (TBI). The “mystical” experience elicited by ibogaine, which is characterized by feelings of awe, selflessness, and transcendence, is correlated with improvements in PTSD symptoms. Mystical experiences with other psychedelics are associated with acute decreases in the activity and connectivity of the default mode network (DMN), which mediates self-related cognition. However, the dynamic effects of ibogaine on the DMN have not yet been studied. At baseline, immediately (3-4 days) after ibogaine, and one month after ibogaine, we acquired resting-state functional magnetic resonance imaging data in an open-label, observational trial of magnesium-ibogaine treatment for 30 U.S. veterans with TBI. Magnesium-ibogaine did not significantly alter static DMN connectivity at either the immediate-post or one-month timepoint. Since static measures cannot capture time-evolving changes in connectivity, we next used Hidden Markov Models (HMM) to measure the post-acute dynamics of DMN activity. Magnesium-ibogaine was associated with significant, sustained decreases in the switching rate (i.e., increases in the duration of) a dynamic DMN substate, which was significantly correlated with the mean score on the Revised Mystical Experience Questionnaire and clinical improvements at one month-post treatment. This DMN substate exhibited a lateral-medial spatial gradient, which was significantly associated with a gradient of externally oriented (i.e., directed to the environment) to internally oriented (i.e., self-related) perception and cognition. Taken together, our results indicate that magnesium-ibogaine alters specific dynamic substates of the DMN, which correlate with its subjective and therapeutic effects.

## Section 1. Introduction

Psychedelics are hallucinogenic compounds that have recently shown promise for treating a wide range of psychiatric and neurological conditions, including post-traumatic stress disorder (PTSD), depression, anxiety, substance use disorders, and traumatic brain injury (TBI).^1–10^ A number of studies have implicated the default mode network (DMN) in the neural mechanisms of psychedelics, demonstrating that they acutely disrupt within-DMN functional connectivity (FC) and activity and increase functional coupling between the DMN and other canonical resting-state networks.^11–16^ (However, a recent meta-analysis found that decreases in within-DMN FC do not always occur on psychedelics.^17^) Located in regions including the medial prefrontal cortex, posterior cingulate cortex, and angular gyrus, the DMN supports self-oriented cognition, autobiographical memory, and mind-wandering.^11,18,19^ Its activity is robustly anticorrelated with the task-positive network of the brain, which is active whenever the brain is engaged in tasks involving the external environment; therefore, the core hubs of the DMN are responsible for internally generated, self-related cognition and perception, though other regions in the DMN may encode more externally-oriented functions.^20–22^

Several psychedelics are often characterized by “mystical” experiences in which the participant’s sense of self dissolves, leading to a feeling of transcendence and unity with the universe.^23–25^ Therefore, the mystical experience may be associated with decreases in within-DMN connectivity; as the self diminishes, so does the DMN, according to certain accounts.^11,26^ In one prominent model named “relaxed beliefs under psychedelics” (REBUS), DMN disruption is understood as a central mechanism of the therapeutic effects of psychedelics.^27^ Other interventions for depression, including transcranial magnetic stimulation, also decrease DMN hyperconnectivity, which may be related to spontaneous experiences of mindfulness (and perhaps mysticism) that occur after treatment.^28–30^

Recently, ibogaine, an atypical psychedelic derived from the Central West African shrub *Tabernanthe iboga*, has demonstrated preliminary efficacy at treating a range of psychiatric conditions. In an observational study of 30 Special Operations Forces veterans with TBI and diverse neuropsychiatric sequelae, a single treatment with the Magnesium-Ibogaine: the Stanford Traumatic Injury to the CNS (MISTIC) protocol was associated with dramatic reductions in symptoms of PTSD, depression, anxiety, and functional disability, with remission rates exceeding 83% at one month across multiple symptom domains.^31^ (Participants were treated with magnesium both before and after the ibogaine dose to mitigate the cardiotoxic effects of ibogaine.) Improvements in symptoms were highly durable, lasting up to one year after treatment.^32^ Qualitative investigation of the same cohort found that the psychological process resembled a form of “auto-therapy,” i.e., a self-directed confrontation with and reprocessing of traumatic material, and that the intensity of the mystical experience during ibogaine treatment correlated with greater reductions in PTSD, depression, and anxiety symptoms at one month.^33,34^ Unlike classic psychedelics like psilocybin and lysergic acid diethylamide (LSD), which more selectively bind to serotonin-2A (5-HT_2A_) receptors, ibogaine has a very complex pharmacology, also acting on the kappa-opioid receptor, N-methyl-D-aspartate (NMDA) receptor, serotonin and norepinephrine transporters, and several other sites.^35–37^

A recent functional magnetic resonance imaging (fMRI) study of the same cohort in the MISTIC trial found that ibogaine altered FC between specific DMN regions and nodes in other brain networks, but these changes in connectivity did not survive multiple comparisons correction.^38^ Both this study and the psychedelic neuroimaging literature more broadly focused on *static* FC measures, which average connectivity across the entire scan and treat the DMN as a temporally uniform entity. In reality, the DMN cycles through distinct states whose temporal dynamics — how long each state persists and how readily the brain transitions between them — carry information that static measures cannot capture.^39–42^

Here, we address these gaps by measuring both static and dynamic FC in resting-state fMRI data collected from the MISTIC cohort^31^ at baseline (BL; 2-3 days before ibogaine), immediately post-treatment (IP; 3-4 days after ibogaine), and one month post-treatment (1M). Whereas the prior fMRI study on this cohort examined static FC by conducting pairwise correlations between regions of interest across the whole brain, here we averaged correlations belonging to the DMN and other networks.^38^ To measure dynamics, we applied Hidden Markov Models (HMMs), a well-established technique in the neuroimaging literature.^43–48^ We found that ibogaine did not significantly alter within-DMN FC, but did significantly alter a particular dynamic substate, whose duration correlated with mystical experiences and clinical outcomes.

## Section 2. Methods

### Section 2.1. Participants and treatments

The data were acquired as part of the previous MISTIC study.^31^ Details are summarized in **Supplementary Methods 1-2**. In brief, 30 male veterans with mild-to-moderate TBI received a single dose of ibogaine (12.1 ± 1.2 mg/kg) and were pre- and post-treated with intravenous magnesium sulfate (1 g) to mitigate cardiovascular risks, particularly QT interval prolongation.

### Section 2.2. Subjective and clinical measures

Mystical experiences on ibogaine were measured retrospectively at the IP timepoint with the Revised Mystical Experience Questionnaire (MEQ-30).^49^ Responses to the questions were averaged into one aggregate mean score. Additionally, various clinical measures, including the Clinician-Administered PTSD Scale for DSM-5 (CAPS-5), Montgomery-Asberg Depression Rating Scale (MADRS), Structured Interview Guide for the Hamilton Anxiety Scale (HAM-A), and World Health Organization Disability Assessment Schedule 2.0 (WHODAS-2.0, a measure of functional disability), were administered at baseline, IP, and 1M and four long-term timepoints, which included 3 months-post, 6 months-post, 9 months-post, and 12 months-post.^32^ Number of TBIs and blast exposure, assessed with the Boston Assessment of TBI - Lifetime (BATL), were measured at baseline. Details on each of these measures are provided in the previous MISTIC study, as well as a separate study reporting the long-term follow-up data.^31,32^

### Section 2.3. fMRI data acquisition and preprocessing

fMRI data were acquired at three timepoints: baseline (*n* = 29), IP (*n* = 28), and 1M (*n* = 25). Data were missing from some participants because of failure to meet MRI safety criteria or other logistical issues. fMRI data acquisition and preprocessing were previously reported^50^ and are summarized in **Supplementary Methods 3.**

### Section 2.4. Data analysis

A list of primary vs. exploratory analyses is provided in **Table 1**.

**Table 1.** Primary vs. exploratory analyses in this study. Generally speaking, correlations between static/dynamic measures of the DMN and MEQ scores were primary, since there is more evidence linking mystical experiences and psychedelic-induced changes in DMN activity or connectivity.

| Analysis | Primary or Exploratory? |
| --- | --- |
| Static within-DMN FC | Primary |
| Static within-network FC (of networks besides the DMN) | Exploratory |
| Static between-network FC | Exploratory |
| Correlation between static within-DMN FC and MEQ | Primary |
| Correlation between static within-DMN FC and clinical outcomes | Exploratory |
| Dynamic DMN substate switching rate | Primary |
| Dynamic DMN substate fractional occupancy | Primary |
| Correlation between dynamic DMN substate switching rate and MEQ | Primary |
| Correlation between dynamic DMN substate switching rate and clinical outcomes (including 12 month follow-up) | Exploratory |
| Correlation between dynamic DMN substate topography and internal-external gradient | Exploratory |

#### Section 2.4.1. Static FC

fMRI data were parcellated with the Schaefer-200 atlas, which is itself divided into the seven Yeo networks: visual network (VN), somatomotor network (SMN), dorsal attention network (DAN), salience network (SN), limbic network (LN), central executive network (CEN), and default mode network (DMN).^51,52^ We then measured the Fisher-transformed Pearson correlation of the FC between all pairs of regions. For static within-network FC, these correlations were averaged across pairs of regions that belonged to the same network. For static between-network FC, correlations were averaged across pairs of regions that belonged to different networks. A linear mixed-effects model (LME) was used to determine statistical significance, with timepoint as a fixed effect and subject as a random effect (FC ∼ Timepoint + (1 | Subject)). *p*-values for timepoint contrasts were corrected for multiple comparisons across timepoints and networks using the Benjamini-Hochberg false discovery rate (FDR) procedure.

Pearson correlations were conducted between MEQ-30 ratings and changes in static within-DMN FC at the IP and 1M timepoints. (The data for all correlations in this study generally met the assumptions required for Pearson correlations, including linearity, lack of extreme outliers, independence of observations, and lack of severe skew.) FDR was used to correct for multiple comparisons across the two timepoints. Exploratory Pearson correlations were also conducted between clinical measures - total scores on the CAPS-5, MADRS, HAM-A, and WHODAS-2.0 scales at IP and 1M - and static within-DMN FC. Due to the exploratory nature of these correlations, only uncorrected *p*-values are reported.

#### Section 2.4.2. Dynamic FC analysis

Hidden Markov Models (HMMs) are probabilistic models that represent a timeseries as a sequence of discrete, recurring latent states, each associated with a characteristic pattern of observed activity.^53,54^ The transitions between states are governed by a Markov process in which the state at a given timepoint depends only on the state at the preceding timepoint. In applications to fMRI and other human brain data, the observed regional timeseries are modeled as ‘emissions’ from these hidden brain states.^43–48^ Different types of HMMs use different distributions to describe the probability of the observed timeseries given the hidden state, with the Gaussian distribution being the most common.^45,46,48^ This framework allows the data to be decomposed into temporally dynamic brain states that can recur across time and across participants, while also estimating the probability of transitioning from one state to another. In contrast to static FC analyses, HMMs explicitly capture moment-to-moment changes in large-scale brain dynamics.

The Schaefer-200 atlas was restricted to the 37 parcels assigned to the DMN. Using the GLHMM toolbox in Python,^46^ Gaussian HMMs with full covariance ranging from *K* = 2 to *K* = 10 states were fit to the concatenated DMN parcel timeseries across baseline, IP, and 1M sessions, while preserving session boundaries. Prior to fitting, parcel timeseries were *z*-scored within sessions. *K* was selected using the elbow of the free-energy curve. For each DMN substate, we quantified fractional occupancy and switching rate using GLHMM outputs. Fractional occupancy was defined as the proportion of time spent in a DMN substate within a session, and switching rate as the rate of transitions between DMN substates within a session. Timepoint effects of summary metrics were tested using LMEs with timepoint as a fixed effect and subject as a random intercept (Summary Metric ∼ Timepoint + (1 | Subject)); post-baseline contrasts were corrected for multiple comparisons using FDR across DMN substate-by-timepoint tests. To ensure that head motion (measured as mean framewise displacement [FD]), age, and number of TBIs did not account for the results, the LME was rerun with these three covariates added as fixed effects (Summary Metric ∼ Timepoint + FD + Age + #TBIs + (1 | Subject)). Pearson correlations were also performed between summary metrics and mean FD at each timepoint.

Pearson correlations were conducted between MEQ-30 ratings and session-normalized changes in switching rate of DMN substate 3 at the IP and 1M timepoints. Note that we focused on the switching rate of DMN substate 3 because it was the only summary metric that exhibited significant changes across both timepoints (see *Section 3.2*). For session normalization, the switching rate of DMN substate 3 per subject per session was divided by the mean of the switching rate across DMN substates for that subject and session. FDR was used to correct for multiple comparisons across the two timepoints. Exploratory Pearson correlations were also conducted between clinical measures - total scores on the CAPS-5, MADRS, HAM-A, and WHODAS-2.0 scales - and switching rate of DMN substate 3. To reduce the number of multiple comparisons, clinical data from only one long-term follow-up visit, i.e., 12 months-post (12M), were correlated with summary metrics. In total, FDR was used to correct for 12 comparisons (3 timepoints [IP, 1M, 12M] x 4 clinical outcomes).

Whole-brain HMMs were also fit to the full Schaefer-200 parcel time series. For these analyses, parcel timeseries were mean-centered across regions at each timepoint to reduce global variance, then standardized within-session and reduced to 15 dimensions using principal component analysis.

Gaussian HMMs with full covariance shared across whole-brain states, ranging from *K* = 2 to *K* = 10 whole-brain states, were trained. *K* was once again selected using the elbow of the free-energy curve. Fractional occupancy and switching rate were then compared across timepoints using the same mixed-effects framework described above.

#### Section 2.4.3. Gradient of internal to external perception

Mystical experiences are characterized by a feeling of oneness with the environment, or a dissolution of the boundaries between the internal and external worlds. We therefore determined whether DMN substates exhibited increased activation in DMN regions involved in externally oriented cognition and decreased activation in DMN regions involved in internally oriented cognition. To do this, we embedded terms in the Cognitive Atlas, a set of words and phrases describing various cognitive phenomena (e.g., “action perception,” “motor planning,” “agency”), along a semantic axis representing external cognition on one end and internal cognition on the other.^55^ A set of eight Cognitive Atlas terms were chosen as anchors for the internal pole of the axis: ‘autobiographical memory’, ‘self monitoring’, ‘introspection’, ‘interoception’, ‘internalizing’, ‘mental imagery’, ‘imagination’, and ‘episodic memory’. Eight separate Cognitive Atlas terms were chosen as anchors for the external pole of the axis: ‘visual attention’, ‘selective attention’, ‘action perception’, ‘motor control’, ‘visual object recognition’, ‘auditory perception’, ‘response selection’, and ‘attention.’ All Cognitive Atlas terms were then represented as vectors in a Term Frequency-Inverse Document Frequency (TF-IDF) matrix, which assigns weights to words based on how strongly they are related to a Cognitive Atlas term.^56^ This matrix is then reduced to 100 dimensions via singular value decomposition (SVD). We averaged the vector embeddings of the anchor terms in this 100-dimensional space, yielding centroids corresponding to the two poles of the internal-external semantic axis. Then, we computed the cosine similarity of the embedding of each Cognitive Atlas term to the ‘internal’ and ‘external’ centroids, thereby scoring each term along the internal-external semantic axis. Positive scores indicated that a term is more related to externally oriented cognition, whereas negative scores indicated that a term is more related to internally oriented cognition.

To relate the internal-external semantic axis to DMN regions, we leveraged prior applications of Neurosynth to identify patterns of brain activity encoding each of the Cognitive Atlas terms. Neurosynth is a tool that automatically performs meta-analyses on fMRI papers about various topics, generating an aggregate brain activation map for each topic.^57^ Previous studies have applied Neurosynth to Cognitive Atlas terms, resulting in a collection of brain activation maps for each term (identifiers.org/neurovault.collection:1274).^58^ Each of these maps was parcellated to the 37 DMN regions in the Schaefer-200 atlas. Maps corresponding to terms with the top 10% score on the internal-external semantic axis (i.e., the most ‘external’ terms) were averaged, and the same was done for the terms with the bottom 10% score. (Note that our result remained significant when the selection quantile was set to 15%, but not at 5% or 20%.) The difference between these two averages was then computed for each DMN region, resulting in an ‘internal-external gradient’ across DMN areas.

Finally, we performed Pearson correlations between the DMN substate means and this internal-external gradient. Statistical significance of the correlation was assessed using a null model that preserves spatial autocorrelation, as implemented in neuromaps.^59^ A custom volumetric parcellation containing the 37 DMN parcels was resampled to MNI152 2mm space, and Moran spectral randomization was used to generate surrogate maps with matched spatial autocorrelation structure.

## Section 3. Results

### Section 3.1. Static FC analysis

A schematic of the study design is shown in **Figure 1**. Ibogaine did not significantly alter static within-DMN FC at either IP (β = −0.016, SE = 0.012, *p*_FDR_ = 0.438) or 1M (β = −0.028, SE = 0.012, *p*_FDR_ = 0.175) (**Figure 2**). The only significant change in static within-network FC occurred in the salience network at IP (β = −0.064, SE = 0.020, *p*_FDR_ = 0.016), but not at 1M. There were also no significant differences in between-network connectivity (**Supplementary Table 1**).

**Figure 1.**
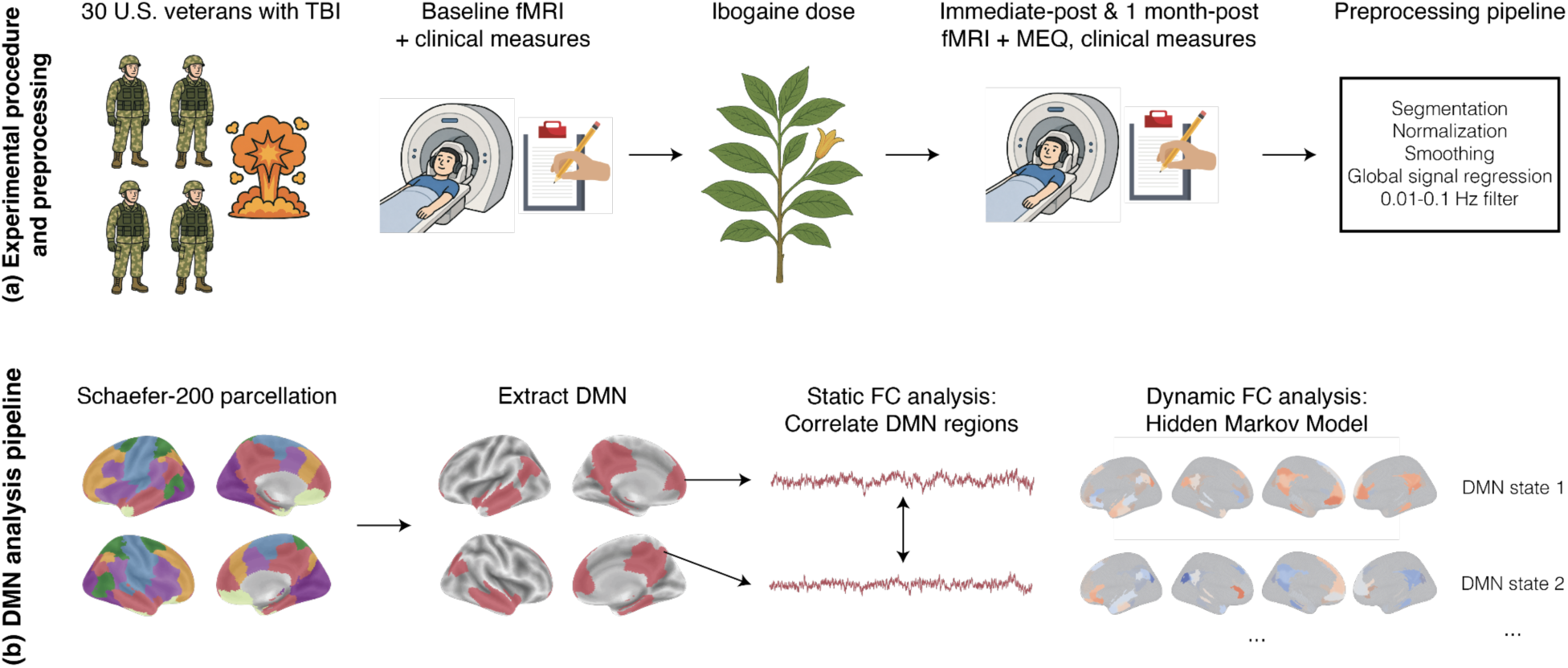
Overview. (a) A single dose of ibogaine (mean dose = 12.1 ± 1.2 mg per kg) was administered to 30 U.S. Special Operations Forces veterans with mild-to-moderate traumatic brain injury (TBI) and varying levels of post-traumatic stress disorder (PTSD), depression, anxiety, and alcohol use disorder. Resting-state fMRI and clinical symptoms, such as symptoms of TBI and PTSD, were measured at baseline, immediate-post (3-4 days after), and one month-post ibogaine. Standard preprocessing was performed with fMRIPrep, including global signal regression. Mystical experiences on ibogaine were measured once, at the IP timepoint, with the revised Mystical Experience Questionnaire (MEQ-30). (b) fMRI data was parcellated with the Schaefer-200 parcellation, which is divided into the seven Yeo networks. These include the default mode network (DMN), which comprises 37 regions. For the static FC analysis, Fisher-transformed Pearson correlations were measured between these DMN regions and averaged. For the dynamic FC analysis, a Hidden Markov Model (HMM) was fitted to the DMN timeseries, yielding five DMN substates. Summary metrics of these substates, including switching rate and fractional occupancy, were compared across timepoints. These metrics, as well as static FC, were each correlated with the mean MEQ-30 score and clinical outcomes.

**Figure 2.**
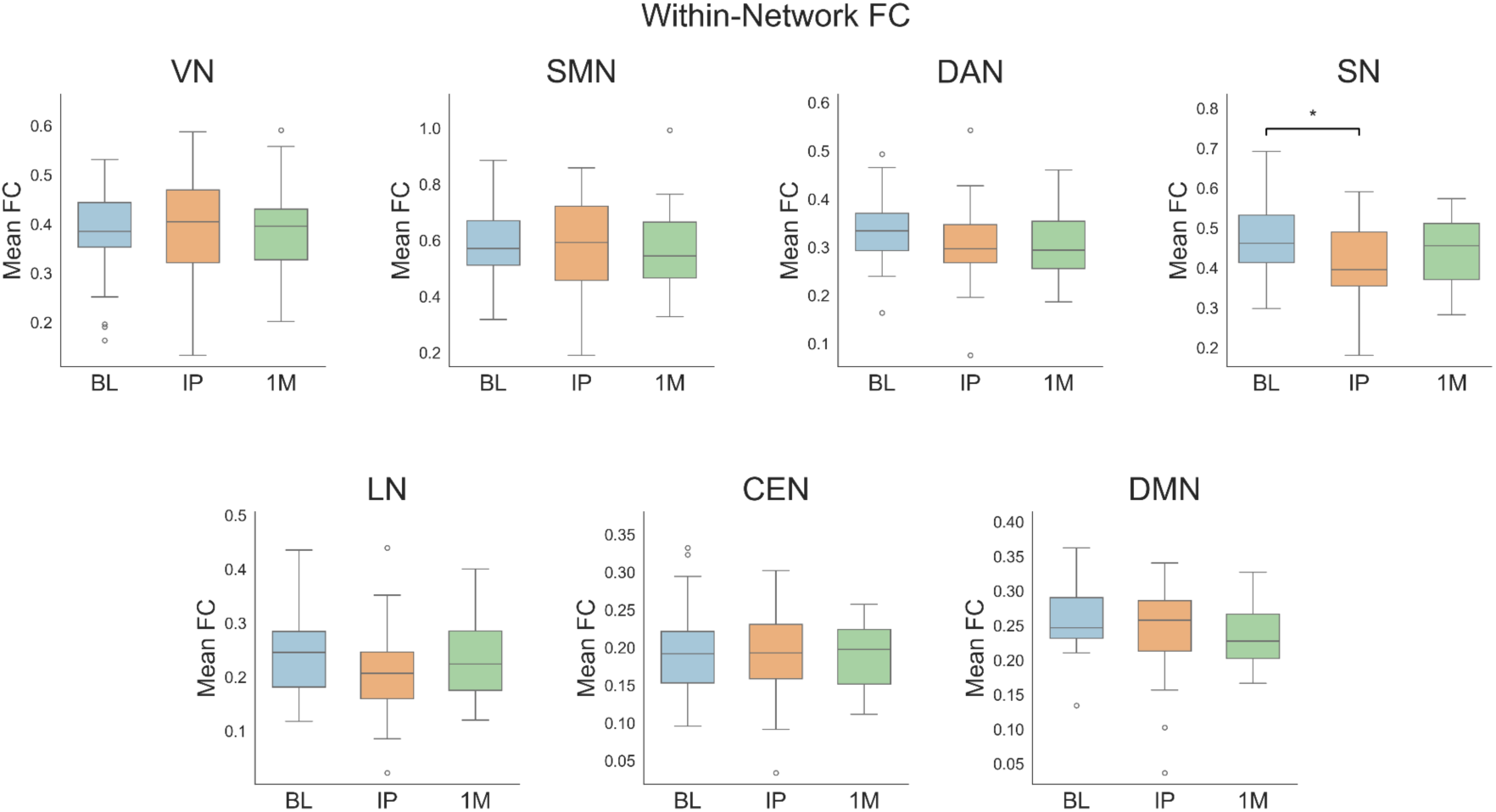
Ibogaine significantly reduces the within-network FC of the salience network, but not any other networks. Within-network FC was computed as the average Fisher-transformed Pearson correlation between the constituent regions of each Yeo network. Statistically significant changes in within-network FC were determined based on a linear mixed-effects model with timepoint as fixed effect, and then FDR correction was used to adjust for multiple comparisons across the seven networks and two timepoints (IP and 1M). The only change that survived multiple comparisons was a decrease in the within-network FC of the salience network (SN) at IP. VN = visual network; SMN = somatomotor network; DAN = dorsal attention network; SN = salience network; LN = limbic network; CEN = central executive network; DMN = default mode network. * 0.01 < *p*_FDR_ < 0.05.

The intensity of the mystical experience, as quantified by the mean score on the MEQ-30, was significantly and positively correlated with changes in within-DMN FC at 1M (*r* = 0.478, *p*_FDR_ = 0.036) (**Figure 3**). The correlation trended towards significance at the IP timepoint (*r* = 0.336, *p*_FDR_ = 0.087). Thus, although within-DMN FC did not exhibit a significant mean shift across the cohort, inter-individual differences in within-DMN FC were positively associated with MEQ. However, there were no significant correlations between any of the clinical outcomes (see Methods section) at either timepoint and within-DMN FC (**Supplementary Figure 1**), or FC within the salience network.

**Figure 3.**
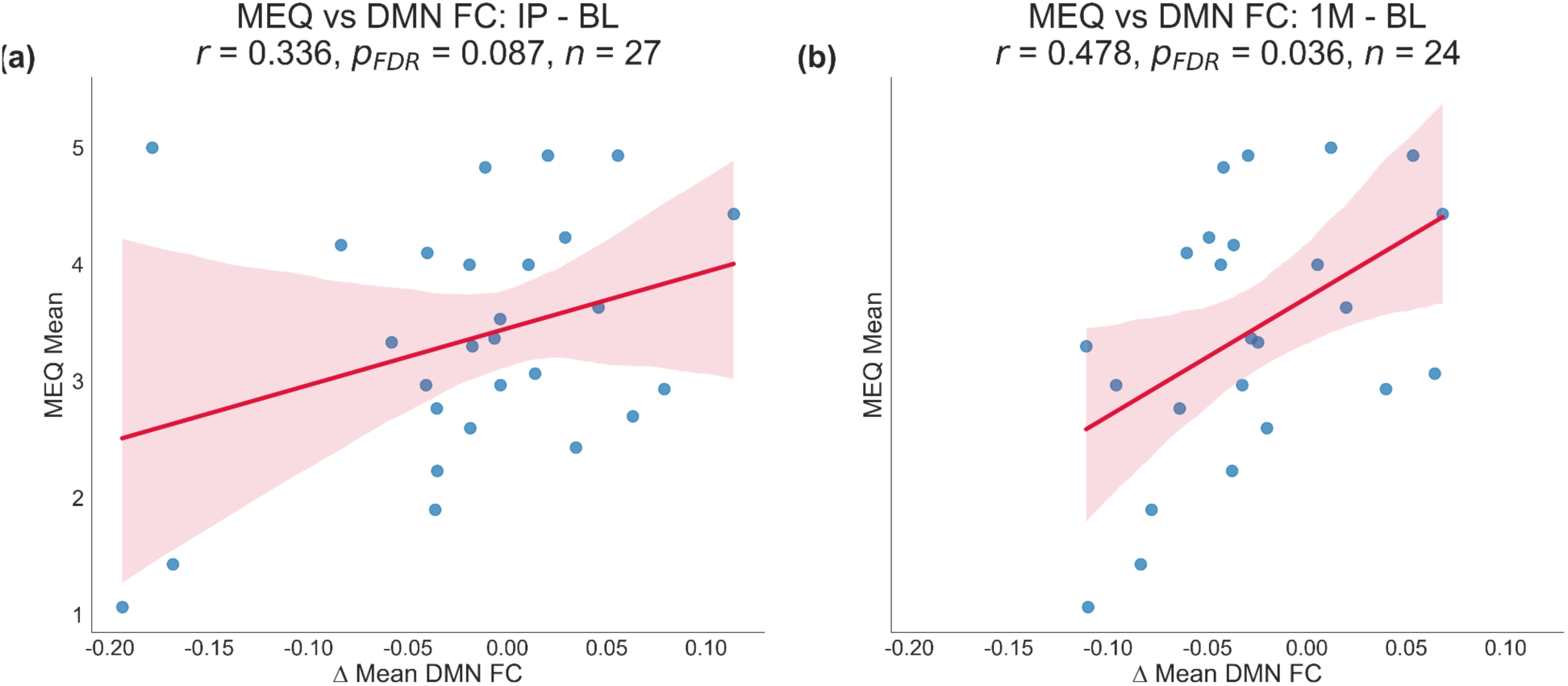
Increases in FC within the DMN one month after ibogaine are significantly correlated with the intensity of the mystical experience. Pearson correlations were conducted between the mean score on the MEQ and the mean within-DMN FC. The MEQ was administered only once, at IP. FDR correction was used to adjust for multiple comparisons across the two timepoints (IP and 1M). Red shaded bands represent the 95% confidence interval for the estimated mean regression line. (a) The positive correlation between increases in within-DMN FC at IP and the total MEQ score trended towards significance. (b) At 1M, there was a significant, positive correlation between increases in within-DMN FC and the total MEQ score.

### Section 3.2. Dynamic FC analysis

Since static FC analysis averages connectivity over the entire scanning session, we next determined whether a dynamical analysis, namely HMMs, could better differentiate DMN activity before and after ibogaine. *K* = 5 was identified as the optimal number of DMN substates based on the free energy of the model (**Supplementary Figure 2**). Each DMN substate was characterized by a different pattern of mean activation across DMN regions (**Figure 4**) and covariance between DMN regions (**Supplementary Figure 3**). While ibogaine was not associated with a significant change in the fractional occupancy of any of the DMN substates (**Supplementary Figure 4**), it was associated with a significant decrease in the switching rate - in other words, it increased the duration - of DMN substate 2 in this analysis at IP (β = −0.010, SE = 0.003, *p*_FDR_ = 0.020) (**Figure 5**). (Note, however, that fractional occupancy and switching rate were generally correlated with each other, across states and timepoints [*r* = 0.59].) Ibogaine was also associated with a significant reduction in the switching rate of DMN substate 3 in this analysis at both IP (β = −0.006, SE = 0.002, *p*_FDR_ = 0.038) and 1M (β = − 0.007, SE = 0.003, *p*_FDR_ = 0.020). The decrease in DMN substate 3 switching rate remained significant after including head motion, age, and number of TBIs as covariates in the LME (IP vs BL: β = −0.007, SE = 0.003, *z* = −2.80, *p*_FDR_ = 0.005; 1M vs BL: β = −0.008, SE = 0.003, *z* = −2.86, *p*_FDR_ = 0.005) (**Supplementary Figure 5a**). Head motion was not correlated with switching rates (**Supplementary Figure 5b**). In summary, the DMN exhibited a significant increase in the duration of a particular dynamic substate, state 3, at both timepoints.

**Figure 4.**
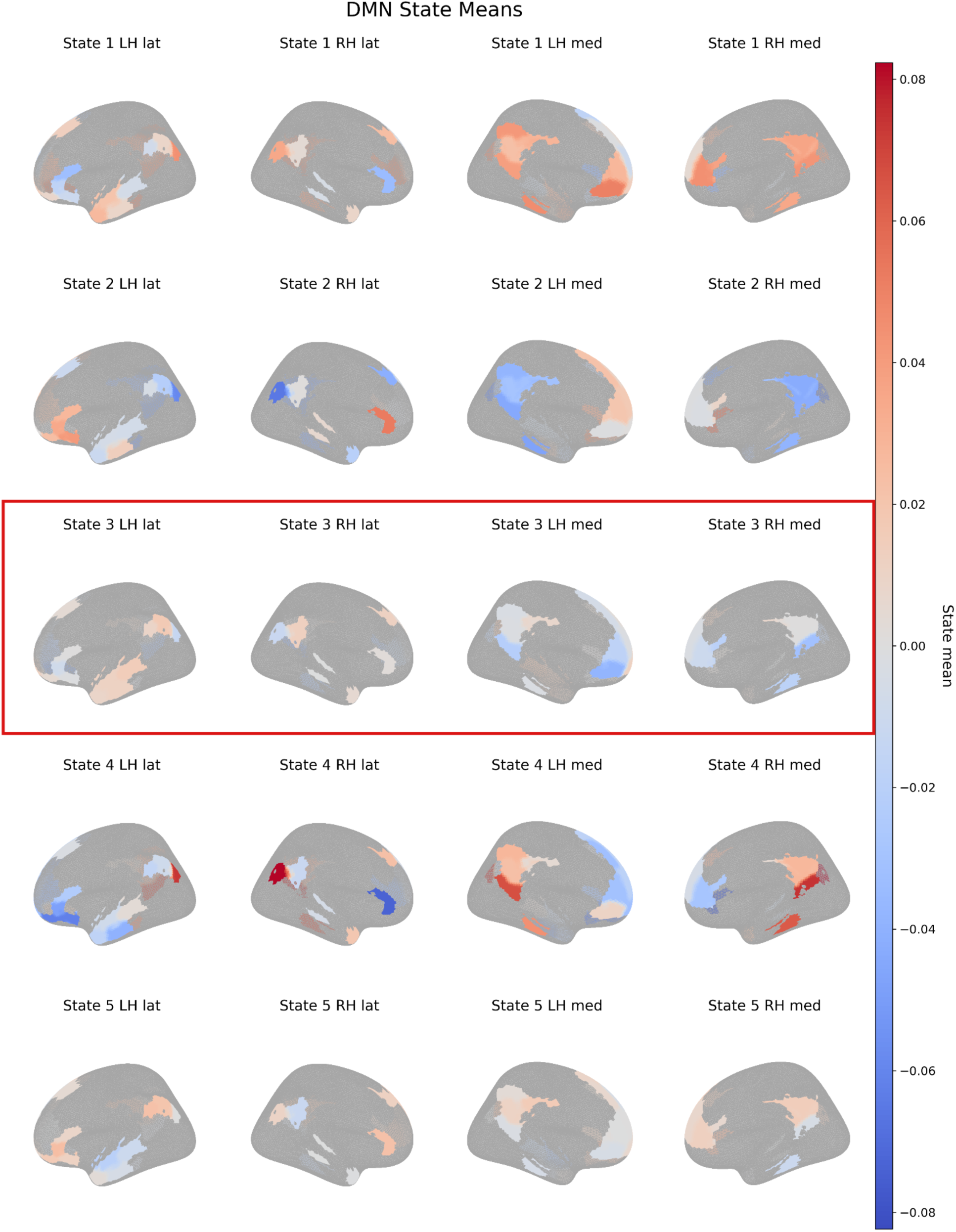
HMM decomposes DMN dynamics into five substates. Each panel shows the mean activity of each substate over the DMN regions, projected onto the cortical surface. Warm colors indicate relatively positive activity and cool colors indicate relatively negative activity. DMN substate 3 is highlighted because it is the only substate with a significant change in switching rate (Figure 5).

**Figure 5.**
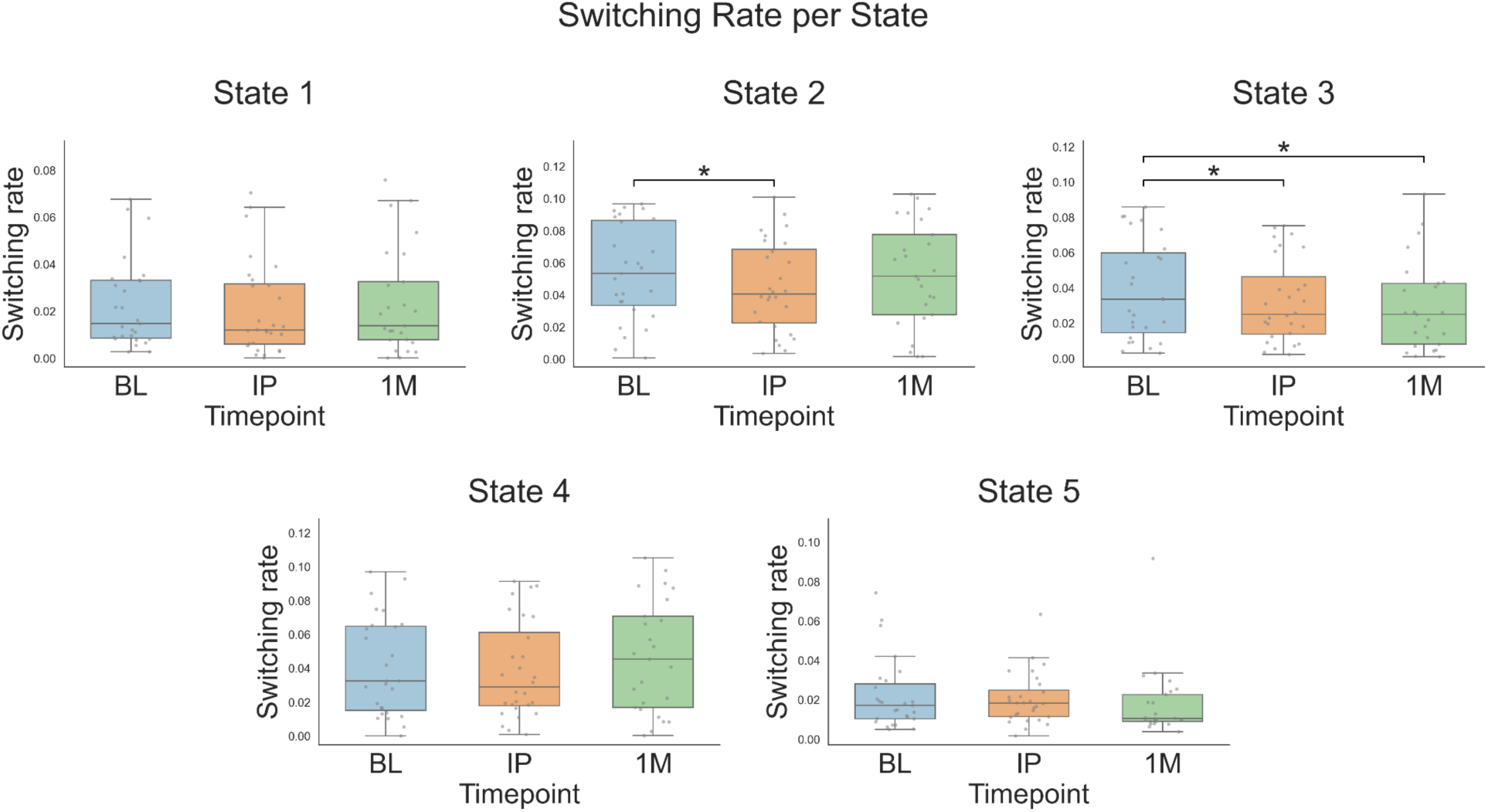
Ibogaine is associated with significant decreases in the switching rate of DMN substate 3. Switching rate is defined as the frequency at which the brain moves from one hidden substate to another over time, and it is inversely related to the duration of a substate. Statistically significant changes in switching rate were determined based on a linear mixed-effects model with timepoint as fixed effect, and then FDR correction was used to adjust for multiple comparisons across the five states and two timepoints (IP and 1M). There was a significant decrease in the switching rate of DMN substate 2 between BL and IP, but not 1M. DMN substate 3 exhibited significant decreases in switching rate at both IP and 1M. In other words, DMN substate 3 exhibited significant increases in duration at both timepoints. * 0.01 < *p*_FDR_ < 0.05.

The change in the session-normalized switching rate of DMN substate 3 at IP was significantly and negatively correlated with the mean MEQ-30 score (*r* = −0.457, *p*_FDR_ = 0.033) (**Figure 6**; correlations between all session-normalized dynamic metrics and all subjective/clinical outcomes are shown in **Supplementary Table 2**). The correlation at 1M was still negative but not significant (*r* = −0.271, *p*_FDR_ = 0.200). The mean activation of DMN substate 3 was higher in lateral than medial regions. This lateral-medial axis was also significantly aligned with a gradient of externally oriented to internally oriented cognition (*r* = 0.346, *p*_spatial_ = 0.045); that is, activation of DMN substate 3 tends to be higher in DMN regions that regulate cognitive processes related to perception of the external environment (e.g., visual attention) (**Figure 7**). Meanwhile, activation of DMN substate 3 is lower in DMN regions involved in mediating internal cognitive phenomena (e.g., autobiographical memory).

**Figure 6.**
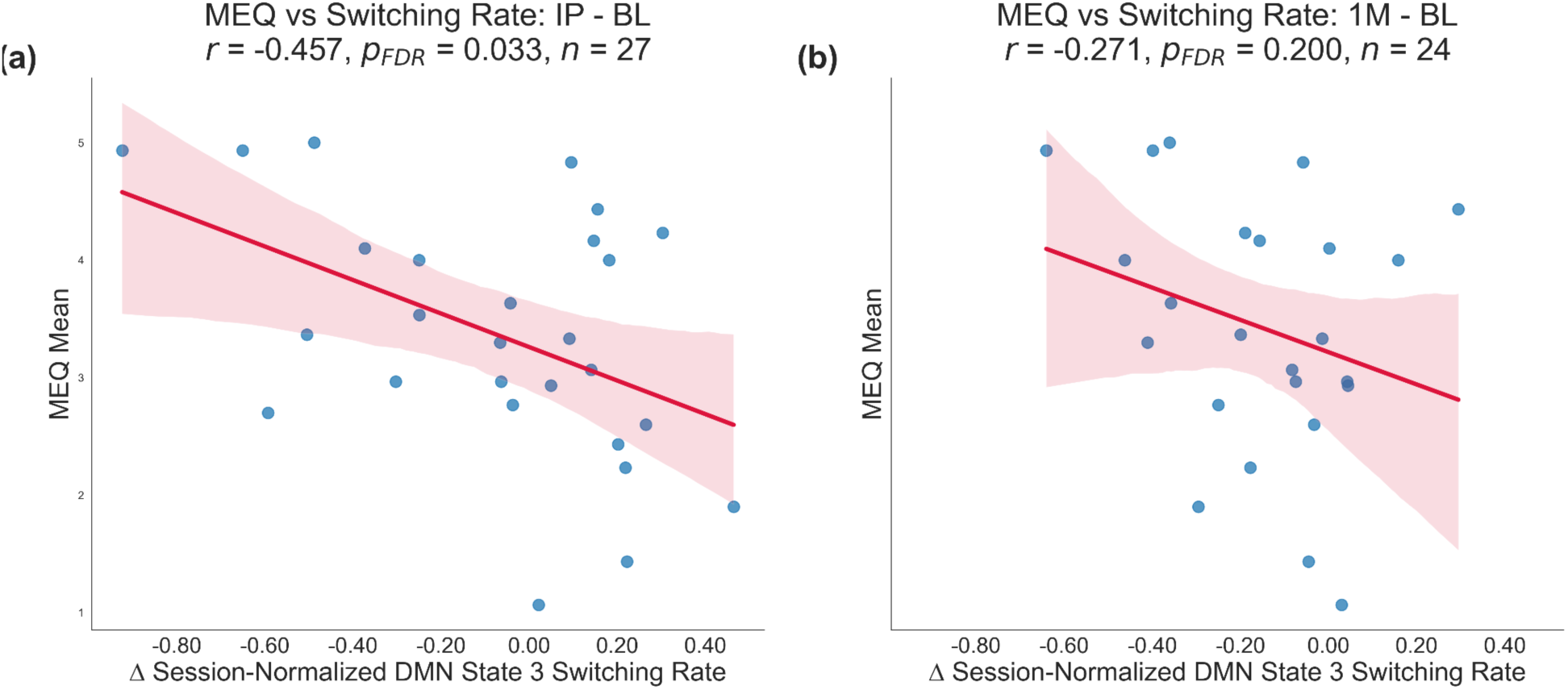
Decreases in the session-normalized switching rate of DMN substate 3 immediately after ibogaine are significantly correlated with the intensity of the mystical experience. To account for inter-individual differences in overall HMM switching propensity, the switching rate of DMN substate 3 per subject per session was divided by the mean switching rate of all five DMN substates for that subject and session. Pearson correlations were conducted between the mean score on the MEQ and the change in the session-normalized switching rate of DMN substate 3 at both timepoints (IP and 1M). FDR correction was used to adjust for multiple comparisons across the two timepoints. Red shaded bands represent the 95% confidence interval for the estimated mean regression line. (a) The change in switching rate at IP is significantly anticorrelated with the mean MEQ score. (b) This correlation is still negative but not significant at 1M.

**Figure 7.**
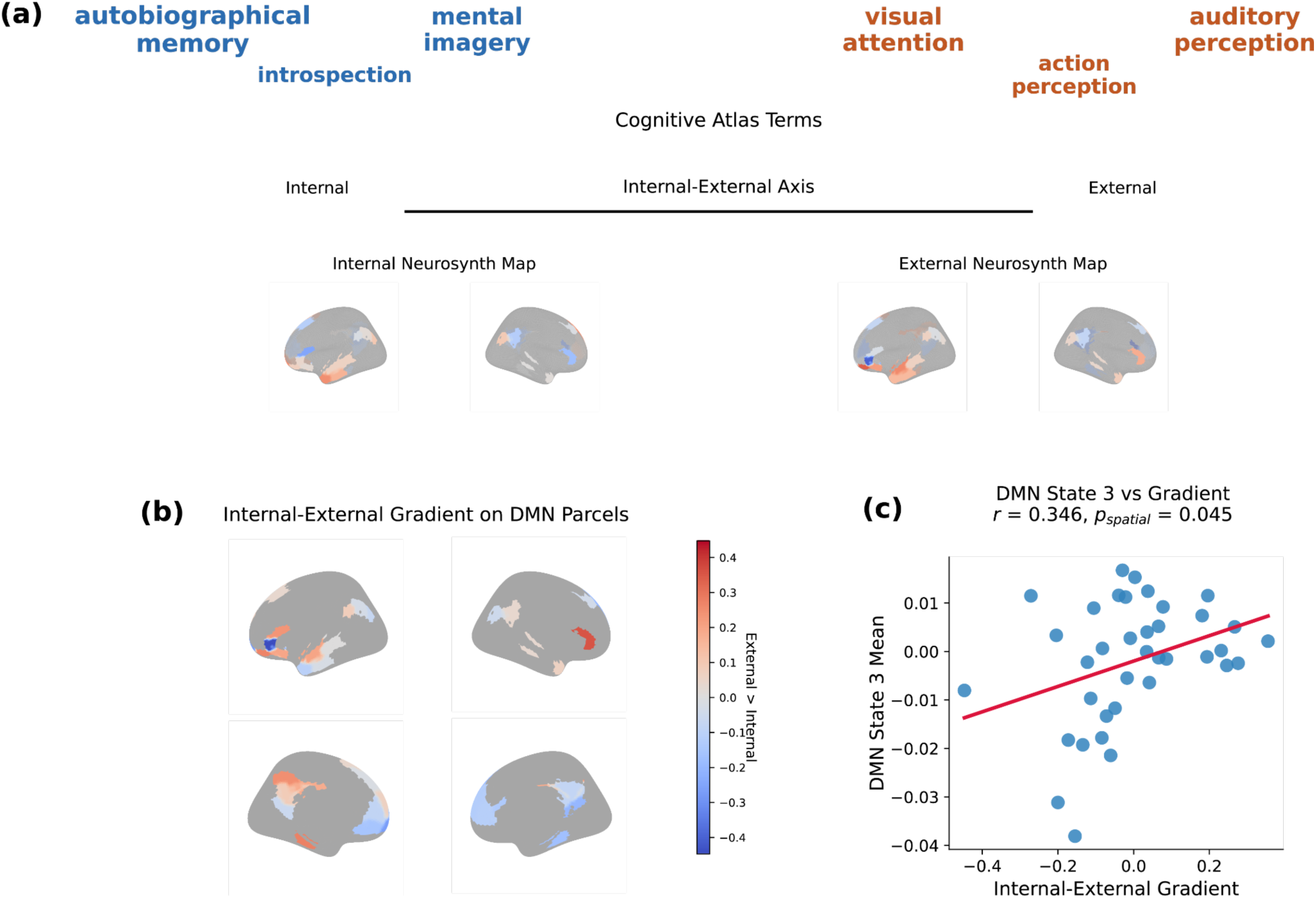
DMN substate 3 means are positively correlated with a gradient of externally to internally oriented perception and cognition. (a) Schematic of the workflow used to construct the gradient. Cognitive Atlas terms were embedded in a latent semantic space and scored along an axis of internally to externally oriented perception and cognition. Specific terms were used as an anchors for ‘internal’ (e.g., autobiographical memory, internal speech, and self knowledge) and for ‘external’ (e.g., visual attention, action perception, auditory perception). Corresponding Neurosynth/NeuroVault activation maps were then aggregated to generate parcel-level internal and external meta-analytic maps. (b) The resulting internal-external gradient projected onto Schaefer-200 DMN parcels, shown on lateral and medial cortical surfaces for both hemispheres. Warmer colors indicate parcels more strongly associated with the external end of the gradient, whereas cooler colors indicate parcels more strongly associated with the internal end. (c) Parcelwise correlation between the DMN internal-external gradient and the mean spatial pattern of DMN substate 3, showing a positive association (*r* = 0.346), consistent with greater positive weights in relatively lateral DMN regions and lower weights in relatively medial DMN regions. This correlation was significant under a spatial autocorrelation-aware null model (*p*_spatial_ = 0.045).

Exploratory analyses revealed that the change in the session-normalized switching rate of DMN substate 3 at the 1M timepoint was significantly and positively correlated with all clinical outcomes at 1M, i.e., scores on the CAPS-5 (*r* = 0.460, *p*_FDR_ = 0.048), MADRS (*r* = 0.567, *p*_FDR_ = 0.015), HAM-A (*r* = 0.700, *p*_FDR_ = 0.001), and WHODAS scales (*r* = 0.548, *p*_FDR_ = 0.018), after FDR correction (**Figure 8**). (Note that the four clinical outcomes may themselves be highly inter-correlated.) In other words, the larger the increase in session-normalized duration of DMN substate 3 (conversely, the greater the decrease in session-normalized switching rate of DMN substate 3) following ibogaine, the greater the clinical improvement on all four scales. None of the other four DMN substates exhibited significant correlations with clinical outcomes at 1M. The change in this switching rate was also significantly and positively correlated with improvements in HAM-A (*r* = 0.627, *p*_FDR_ = 0.015) and WHODAS (*r* = 0.551, *p*_FDR_ = 0.035) scores at 12M (**Supplementary Figure 6**). There was no significant correlation between TBI symptoms at baseline with changes in the session-normalized switching rate of DMN substate 3 at either IP or 1M (number of TBIs: *r*_IP_ = −0.02, *r*_1M_ = 0.01; BATL: *r*_IP_ = −0.16, *r*_1M_ = −0.29).

**Figure 8.**
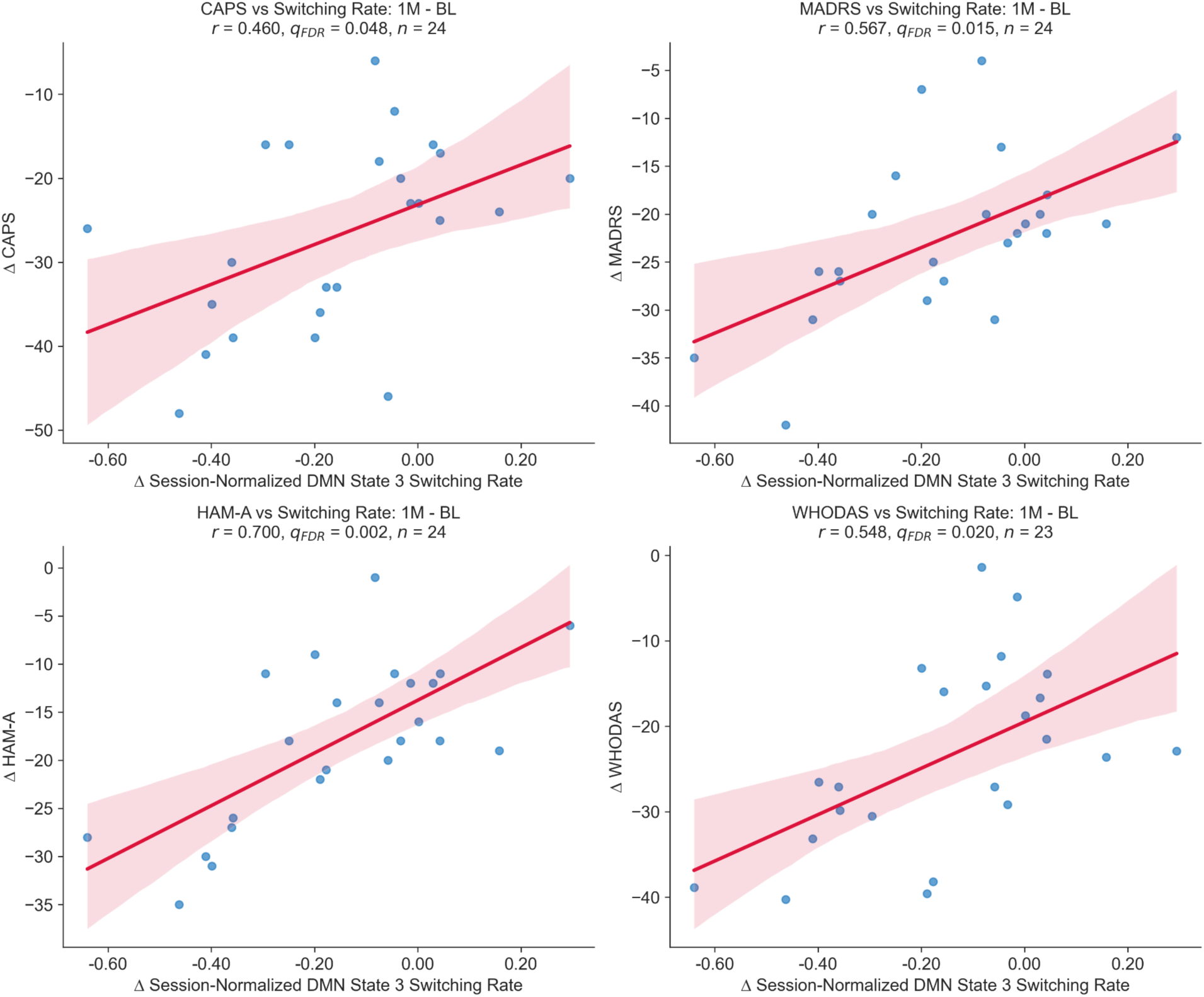
Changes in session-normalized switching rate of DMN substate 3 are significantly correlated with clinical improvements one month after ibogaine. To account for inter-individual differences in overall HMM switching propensity, the switching rate of DMN substate 3 per subject per session was divided by the mean switching rate of all five DMN substates for that subject and session. Pearson correlations were computed between total scores on various clinical scales and session-normalized switching rate of DMN substate 3. These scales included the CAPS-5, which measures PTSD; MADRS, which measures depression; HAM-A, which measures anxiety; and WHODAS, which measures functional disability. Session-normalized switching rate of DMN substate 3 was significantly and positively correlated with all clinical outcomes at the 1M timepoint.

HMMs trained on the whole-brain timeseries revealed states that were aligned with canonical resting-state networks, including the DMN (**Supplementary Figure 7**). However, none of these whole-brain states exhibited significant changes in switching rate at either IP or 1M (**Supplementary Figure 8**). The switching rate of the whole-brain state that most strongly resembled the DMN did not correlate significantly with MEQ-30 mean score or scores on any of the four clinical scales at either IP or 1M (**Supplementary Figure 9-10**).

## Section 4. Discussion

To our knowledge, this is the first study to characterize the post-acute effects of ibogaine on the dynamics of the DMN. Our findings make three central contributions. First, dynamic measures of DMN activity were more sensitive to ibogaine than static measures, both in differentiating pre-from post-treatment brain activity and in correlating with clinically meaningful outcomes. While static measures can provide valuable insights into time-averaged connectivity, dynamic analyses provide complementary information by revealing the evolution of DMN activity over time. Second, we identified a specific dynamical substate of the DMN, state 3 of this analysis, whose switching rate was significantly reduced after ibogaine. This switching rate was correlated with both the intensity of the mystical experience and clinical improvement across multiple symptom domains at one month and at long-term follow-up. Third, this same DMN substate exhibited a lateral-medial spatial gradient, which may be aligned with an axis of externally-to-internally oriented perception and cognition.

### Section 4.1. Dynamic measures are complementary to static measures of ibogaine’s post-acute effects on the DMN

Ibogaine was not associated with significant changes in static within-DMN FC or static between-network FC at either IP or 1M. This null result converges with our group’s prior work on the same cohort: Sridhar and colleagues (2026), using exploratory static FC analyses between pairs of regions, observed widespread changes in static FC across the DMN and other brain networks, yet these did not survive correction for multiple comparisons.^50^ However, the present study is the first to show that static within-DMN FC at 1M was significantly associated with mean scores on the MEQ-30.

The absence of post-acute changes in static within-DMN FC, as well as its positive correlation with mystical experiences, may appear to be in tension with prior literature. Indeed, previous studies have shown that psychedelics reduce FC within the DMN and that mystical experiences are negatively correlated with within-DMN connectivity.^11,60–63^ However, these studies acquired fMRI data *during* the psychedelic experience, whereas the fMRI data in the MISTIC trial was collected 3-4 days and one month *after* ibogaine dosing. (Given the 28- to 49-hour half-life of noribogaine, the primary active metabolite of ibogaine, it is possible that noribogaine was still present in the brain at the IP timepoint.^64^) Furthermore, there is evidence that the post-acute effects of psychedelics on static FC can be the opposite of their acute effects. For example, one day after psilocybin treatment for depression, static within-DMN FC rises, yet it decreases during the treatment.^61,65^ (Note that mystical experiences were measured retrospectively, at the IP timepoint.)

Notably, the one significant change in static FC was not in the DMN but in the salience network, which showed a reduction in within-network static FC at the IP, but not 1M, timepoint. The salience network is a core hub of threat detection, interoceptive awareness, and hyperarousal, and is hyper-connected in patients with PTSD.^66–68^ Its acute disruption following ibogaine may suggest that ibogaine relaxes the hypervigilance that characterizes PTSD, which is supported by the decreases in peak alpha frequency that were observed after ibogaine treatment in electroencephalography (EEG) data from the same veterans.^69–73^ However, salience network FC returned to baseline levels at 1M, so it cannot be a marker of the long-term effects of ibogaine.

By contrast, dynamic FC analysis using HMMs revealed robust, significant effects of ibogaine treatment on the DMN. Ibogaine was associated with significant decreases in the switching rate of DMN substate 3 at both the IP and 1M timepoints, an effect that survived FDR correction across all five DMN substates and two post-treatment timepoints and was robust to inclusion of head motion, age, and number of TBIs as covariates. Crucially, after FDR correction, change in the switching rate of DMN substate 3 was also correlated with the intensity of the mystical experience and with clinical improvements at 1M on the CAPS-5, MADRS, HAM-A, and WHODAS-2.0, as well as with long-term changes in HAM-A and WHODAS-2.0 scores at 12M. Static within-DMN FC, by contrast, was not significantly correlated with any clinical outcome at any timepoint, although it did correlate with the mystical experience at 1M.

Dynamic FC patterns often diverge strongly from static FC, since resting-state FC cycles through different states over time while static measures treat brain networks as if they occupied a single configuration across the entire scanning session^74,75^ Key hubs of the DMN in particular, such as the posterior cingulate cortex, exhibit multiple dynamical co-activation patterns, each one engaging a distinct set of brain regions.^76^ Although HMMs are typically applied to fMRI data of the entire brain, prior work has also fitted HMMs just to the timeseries of DMN regions and recovered discrete DMN connectivity states with distinct dynamical properties.^43,45,48,77^ Most studies of FC in the psychedelic literature have used static measures, but dynamic FC measures are capable of capturing more information.^16,60,78–85^ For instance, only dynamical analyses can reveal whether the ‘repertoire’ of FC states - specific patterns of connectivity between selected regions - expands under psychedelics; static measures only identify a single set of connections over the entire duration of the scan.^83^ As shown in this study, dynamical measures can also indicate changes in the switching rate or duration of certain states, which could occur even when the time-averaged signal remains the same.^84,85^ Post-acute fMRI activity on ibogaine only appears to exhibit significant changes in FC when measured dynamically rather than statically, highlighting the complementary value of dynamic measures.

In summary, static FC appears to capture meaningful individual variability related to mystical experiences, whereas dynamic analyses capture the temporal organization of DMN activity that relates to both mystical experiences and clinical improvement.

### Section 4.2. Substate-specific changes within the DMN

A second contribution of this work is both methodological and conceptual: the prior psychedelic neuroimaging literature has, with few exceptions, treated the DMN as a single, monolithic entity. That is, studies typically report just one within-DMN FC value or a single DMN-component time series; hence, the DMN as a whole goes ‘up’ or ‘down’ in connectivity, which obscures more nuanced, spatially selective changes in FC. In our study, only one of the five DMN substates, state 3, showed a significant change in switching rate both immediately and one month after ibogaine. Were the DMN analyzed only as a single composite signal, the ibogaine-induced change in DMN substate 3 would have been diluted by the unchanged dynamics of the other four DMN substates, plausibly contributing to the null static-FC findings.

The whole-brain HMM analysis reinforces this point. When the HMM was fit to the full 200-parcel time series rather than restricted to DMN parcels, the whole-brain state that best resembled the *entire* DMN did not show significant changes in switching rate after ibogaine, and none correlated with the mystical experience or clinical outcomes. Identifying DMN *substates* with a DMN-restricted HMM was necessary to identify dynamics that differentiated brain activity before and after ibogaine.

The gradient analysis offers a principled explanation of why DMN substate 3, specifically, should be the substate that tracks mystical experience. DMN substate 3 was characterized by relatively higher activity in lateral DMN regions and lower activity in medial DMN regions. While the core, medial hubs of the DMN are associated with internally oriented, self-referential cognition, lateral regions outside these hubs encode different behaviors.^21,86,87^ In particular, more lateral regions, including lateral temporal cortex, the inferior parietal lobule, and middle temporal gyrus, are recruited by more externally directed semantic, attentional, and social tasks, while medial DMN areas deactivate in response to these tasks.^21,22,86–89^ Decreased switching rate of, or increased duration of, DMN substate 3 may therefore imply that ibogaine recruits lateral DMN regions that are involved in externally oriented cognitive processes (e.g., visual attention, action perception, auditory perception) while reducing activation of medial DMN regions that mediate internally oriented processes (e.g., autobiographical memory, introspection, mental imagery). This reconfiguration of the DMN is consistent with the phenomenology of mystical experiences, in which self-focused awareness recedes and people feel “one” with the surrounding environment.^88–90^ Although the acute effects of ibogaine involve internal, dreamlike experiences, the integration in the days and months afterwards, when we recorded fMRI, often leads to heightened awareness of and re-engagement with the external environment.^91,92^ However, the involvement of lateral DMN regions in externally-oriented functions and medial DMN regions in internally-oriented processes was not directly verified by experiment in this study. Instead, the gradient analysis relied on Neurosynth, which performs a meta-analysis of the literature to determine brain regions that are correlated with various behaviors. The functional relevance of the medial-lateral gradient of DMN substate 3 requires further validation.

Beyond its apparent association with the mystical experience, the change in switching rate of DMN substate 3 was correlated with clinical improvement across multiple symptom domains: PTSD, depression, anxiety, and functional disability. Critically, no such associations were observed for static within-DMN FC, nor for any of the other four DMN substates or for the DMN-resembling state of the whole-brain model. In other words, the clinical relevance of ibogaine’s post-acute effects on the DMN was specific to the temporal dynamics of a single DMN substate, rather than the network’s time-averaged connectivity or its dynamics as a whole. DMN substate 3’s correlation with both the mystical experience and clinical outcomes is consistent with prior work showing that the subjective effects of psychedelics are related to their therapeutic benefits.^34,93–96^ That is, the more intense the subjective experience, the greater the clinical improvement. Although PTSD, depression, and anxiety are typically associated with aberrations in static within-DMN FC, our results indicate that increases in the duration of a particular DMN substate are more predictive of symptom improvement after ibogaine treatment than static measures averaged across the entire DMN.^97–103^ Overall, the substate-specific dynamics of the DMN appear to be a robust neural correlate of the therapeutic and mystical effects of ibogaine.

### Section 4.3. Limitations and future directions

Several limitations should be acknowledged. First, ibogaine treatment was open-label; future studies should consider double-blinded or dose-controlled study designs. Currently, we cannot disentangle the effects of ibogaine from expectancy, set, and setting, especially in light of the fact that the ibogaine treatment center offered other wellness activities such as sweat lodge, massage, yoga, reiki, breathwork and meditation. Second, our sample was all-male and almost completely White. Findings may not generalize to women, to non-White populations, to non-veteran patients, or even other types of veterans (including non-SOVs). Thirdly, this study did not consider other methods of measuring DMN dynamics, such as leading eigenvector dynamics analysis, co-activation patterns, or sliding window FC. Fourthly, the gradient analysis is not robust to different selection quantiles (i.e., thresholds at which DMN regions were evaluated for their involvement in external and internal processes) and may not be robust to different anchor terms. Finally, future studies should acquire fMRI data both during and after the ibogaine experience so that the post-acute and acute neural signatures can be directly compared.

## Section 5. Conclusion

In a cohort of veterans with TBI, dynamic measures captured the reorganization of DMN activity after ibogaine treatment. Ibogaine was specifically associated with an increase in the duration of a single DMN substate whose duration correlated with the intensity of the mystical experience and clinical improvement across PTSD, depression, anxiety, and functional disability. These results argue against treating the DMN as a temporally and spatially uniform network in psychedelic neuroimaging, and suggest that the post-acute neural signature of ibogaine treatment may be best resolved at the level of specific dynamical DMN substates.

## Acknowledgments

This work was made possible by the generous donations of Steve and Genevieve Jurvetson, the Effie and Wofford Cain Foundation, Eugene Jhong, the Saisei Foundation, Laura Keller, and by support from and collaboration with Veterans Exploring Treatment Solutions (VETS), Inc., a non-profit organization committed to advancing healthcare options for veterans. The funders played no role in study design, execution, data analysis, or preparation of the manuscript.

Several authors of this manuscript are employed by and receive financial support from the WOMEN Center of Excellence and/or the War Related Illness and Injury Study Center at the Veterans Affairs Palo Alto Healthcare System. The contents do not represent the views of the U.S. Department of Veterans Affairs or the United States Government.

We gratefully acknowledge the late Dr. Nolan Williams, who conceived and designed the study from which these data derive. This manuscript presents a secondary analysis of that work; Dr. Williams passed away prior to its completion and is therefore not included as an author, but the study itself, and the data underlying this analysis, exist because of his scientific vision and leadership.

## Author Contributions

K.S. conceptualized and conducted the dynamic analyses. C.O. conceptualized and conducted the static analyses. K.S. and C.O. co-wrote the manuscript. T.H. and C.D. assisted with conducting some dynamic analyses, including the fitting of the HMM to DMN timeseries. A.A., M.Sr., A.D.G., and S.H. acquired the fMRI data. A.A. and M.Sr preprocessed the fMRI data. K.N.C. performed screening, psychological and cognitive assessments, supervised related scoring and data entry for MISTIC, and led MISTIC study execution at Stanford for all participants. J.N.K. designed, performed and interpreted the statistical analyses of the initial clinical outcomes from MISTIC (e.g., improvements in depression after ibogaine). R.E.B. performed cognitive assessments, scoring of assessments, and clinical data entry for MISTIC. D.M.B. edited the manuscript. J.P.C. assisted with study design and supported MISTIC study execution at Stanford. I.H.K. assisted with MISTIC study design, supported MISTIC study execution at Stanford, and was protocol director for MISTIC at one point in time. R.D.A., M.A., and M.M.A. edited the manuscript. M.Sa and C.R. co-supervised the project. All authors contributed to editing the manuscript.

## Data and code availability

- Data: The deidentified human participant data reported in this study cannot be deposited in a public repository because they contain sensitive clinical and neuroimaging data from a vulnerable veteran population and are subject to Institutional Review Board-mandated controlled access. To request access, please contact the Stanford University Institutional Review Board and the corresponding author (Cammie Rolle) with a research proposal and data security plan consistent with Stanford data-sharing requirements.
- Code: Original code for the manuscript comprises the internal-external gradient analysis, which can be found here: https://github.com/kfshinozuka/ibogaine-DMN. Otherwise, code used existing functions.

## Supplementary Materials

### Supplementary Methods

#### Supplementary Methods 1. Participants

The MISTIC clinical trial was preregistered on ClinicalTrials.gov (identifier: NCT04313712). A total of 34 individuals were screened for participation, of whom 30 met eligibility criteria and completed both pre- and IP (3-4 days after ibogaine) treatment assessments. All participants were male Special Operations Veterans (SOVs) with a documented history of traumatic brain injury (TBI) due to blast exposure, head trauma, or combat. The mean age was 44.9 years (s.d. = 7.5). 28 out of 30 participants were White, which is a limitation of this study; findings may not generalize to non-White participants. Race, age, sex, and gender were self-reported. Participants reported an average of 38.6 (s.d. = 52.4) prior TBIs, ranging in severity from mild (n = 28) to moderate (n = 1) and moderately severe (n = 1), as assessed using the Ohio State University TBI Identification Method. Psychiatric diagnoses at study entry, determined via the Mini International Neuropsychiatric Interview (MINI), included PTSD (n = 23), major depressive disorder (n = 15), anxiety disorder (n = 14), and alcohol use disorder (n = 15). In line with treatment safety protocols, participants discontinued medications with potential for negative interactions with ibogaine, including diuretics, CYP2D6-inhibiting medications, serotonergic medications (that is, any that may increase risk of serotonin syndrome), calcium channel blockers, β-blockers, benzodiazepines, stimulants, corticosteroids and all psychiatric medications.

#### Supplementary Methods 2. Treatment

All participants were referred by the nonprofit organization Veterans Exploring Treatment Solutions, Inc. (VETS), and, prior to referral, had independently arranged to receive ibogaine treatment at Ambio Life Sciences in Mexico. The MISTIC protocol involved oral administration of ibogaine (mean total dose = 12.1 ± 1.2 mg per kg). One to two hours prior to and 12 hours after ibogaine dosing, participants were pre-treated with intravenous magnesium sulfate (1 g) to mitigate cardiovascular risks, particularly Q-T interval prolongation. Prior to dosing, participants underwent medical evaluation and preparatory sessions with a licensed therapist regarding the ibogaine experience. These sessions included intention setting, tools for setting expectations and psychological preparation for the ibogaine experience, including anxiety management. Integration support was made available following treatment, in which therapists helped patients to process emotions and insights from the ibogaine experience. No psychotherapy was provided during the dosing session itself. Because the psychoactive effects of ibogaine can persist for 24-72 hours or longer, participants remained under continuous medical monitoring for 72 hours post-administration.

#### Supplementary Methods 3. fMRI data acquisition and preprocessing

All participants were screened for MRI safety before scanning procedures. Scans were acquired using a 3T GE Discovery MR750 scanner with a 32-channel head-neck imaging coil at the Center for Cognitive and Neurobiological Imaging at Stanford University. Whole-brain structural images were collected using GE’s BRAVO sequence (3-dimensional, T1-weighted [T1w], FOV = 256 x 256 mm, matrix = 256 x 256 voxels, TR = 6.39 ms, TE = 2.62 ms, slice thickness = 0.9 mm, flip angle = 12°).

One whole-brain dual BOLD-ASL scan was also acquired, but is not analyzed in this study. Two 8-minute whole-brain resting-state scans were acquired consecutively (TR = 1300 ms, TE = 30 ms, flip angle = 60°, slice multiband acceleration factor = 6, FOV = 230 x 230 mm, matrix = 128 x 128 voxels). All participants were instructed to keep their heads still and eyes open during the scan. During the resting-state task, they were asked to let their minds wander freely and avoid repetitive thoughts while attending to a white fixation cross on a black screen. Memory foam and inflatable padding were used to restrict head motion. Additionally, participants’ alertness was monitored using in-scanner video cameras.

All fMRI blood-oxygenation-level-dependent (BOLD) data were preprocessed using fMRIPrep 20.2.6,^104,105^ based on Nipype 1.7.0.^106^ Anatomical T1w images were skull stripped and corrected for intensity nonuniformity. Brain tissue segmentation of cerebrospinal fluid (CSF), white matter (WM), and gray matter was then performed on the brain-extracted T1w images. Finally, nonlinear registration was used to perform volume-based spatial normalization to standard spaces (MNI152NLin2009cAsym). The preprocessing steps were performed on both BOLD resting-state runs for each participant. First, a reference volume and its skull-stripped version were generated using a custom methodology of fMRIPrep. This was followed by an in-house pipeline applied, where all data were spatially smoothed (6-mm full width at half maximum Gaussian kernel); nuisance variables from nonbrain tissue signals (CSF, WM), global signal, and 24 motion parameters were regressed from the data; and a temporal bandpass filter (0.01–0.1 Hz) was applied.

### Supplementary Figures and Tables

**Supplementary Figure 1.**
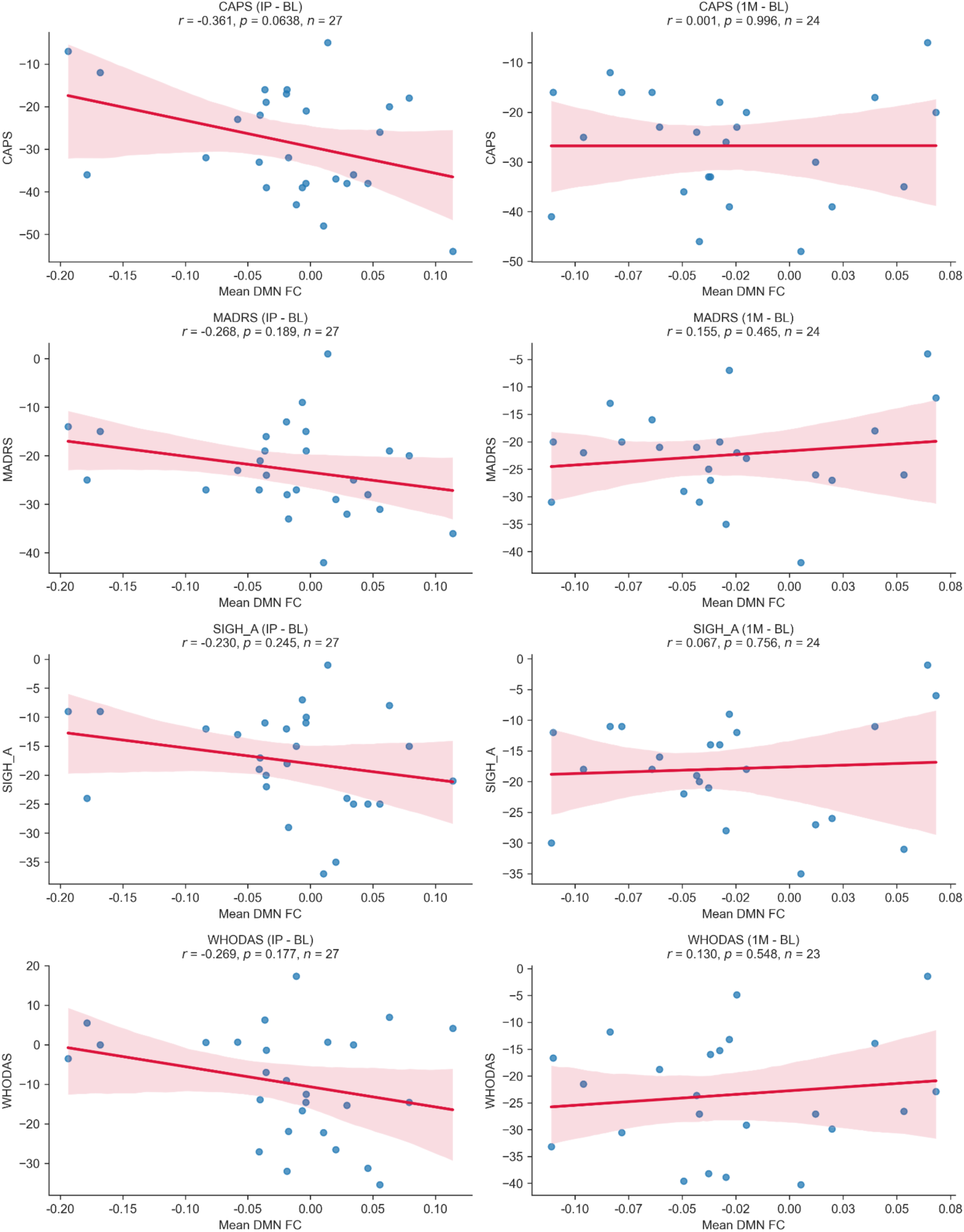
Changes in FC within the DMN are not correlated with clinical outcomes. Pearson correlations were computed between total scores on various clinical scales and within-DMN FC. These scales included the CAPS-5, which measures PTSD; MADRS, which measures depression; HAM-A, which measures anxiety; and WHODAS, which measures functional disability. Due to the exploratory nature of these correlations, only uncorrected *p*-values are reported. None of the uncorrected *p*-values were significant.

**Supplementary Figure 2.**
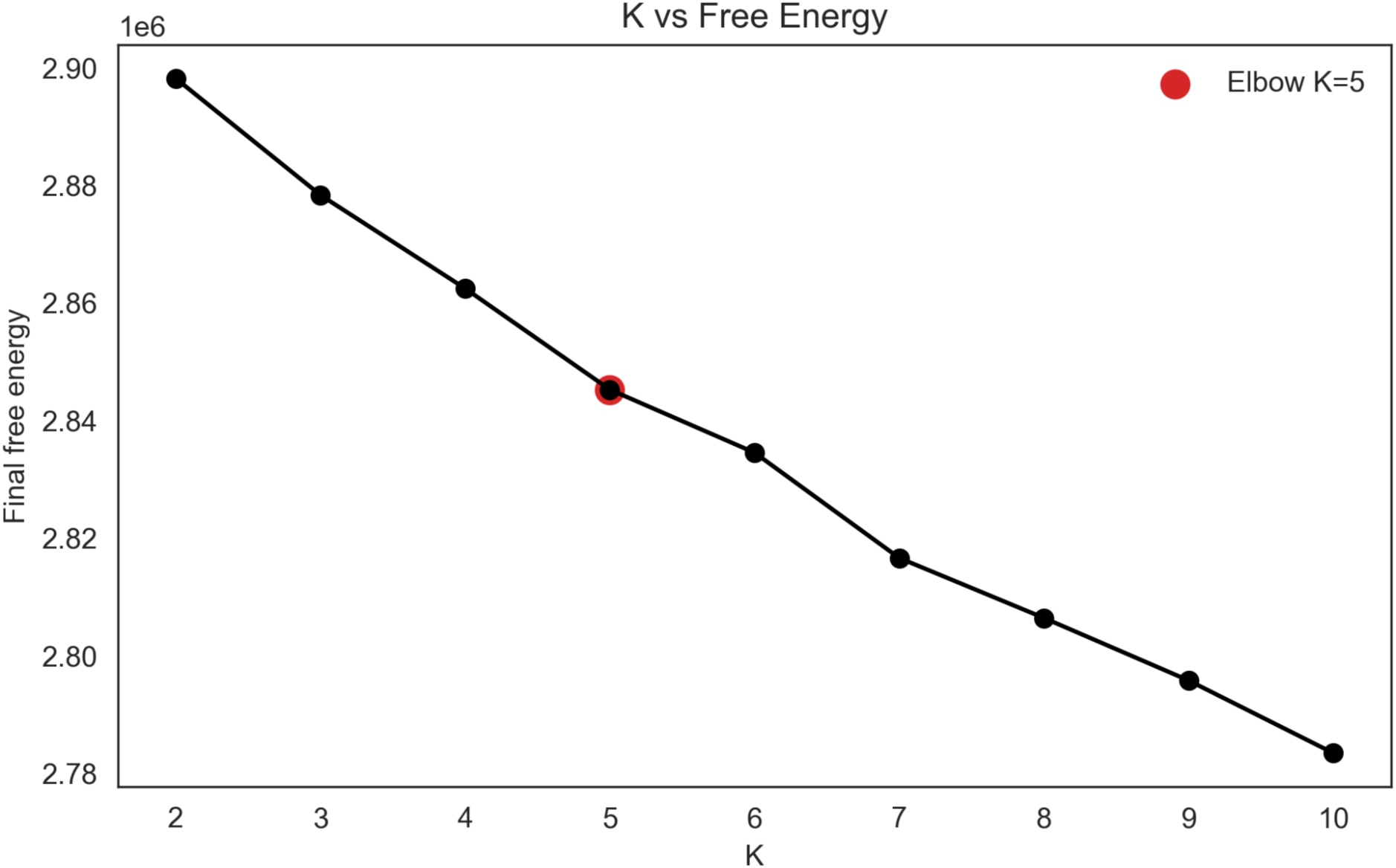
Optimal number of HMM states is *K* = 5. We fitted HMMs ranging from *K* = 2 to *K* = 10 states. For each *K*, we computed the free energy, i.e., how well the HMM explains the observed data while penalizing unnecessary complexity. The optimal *K* was selected based on the first “elbow” of the *K* vs. free energy plot.

**Supplementary Figure 3.**
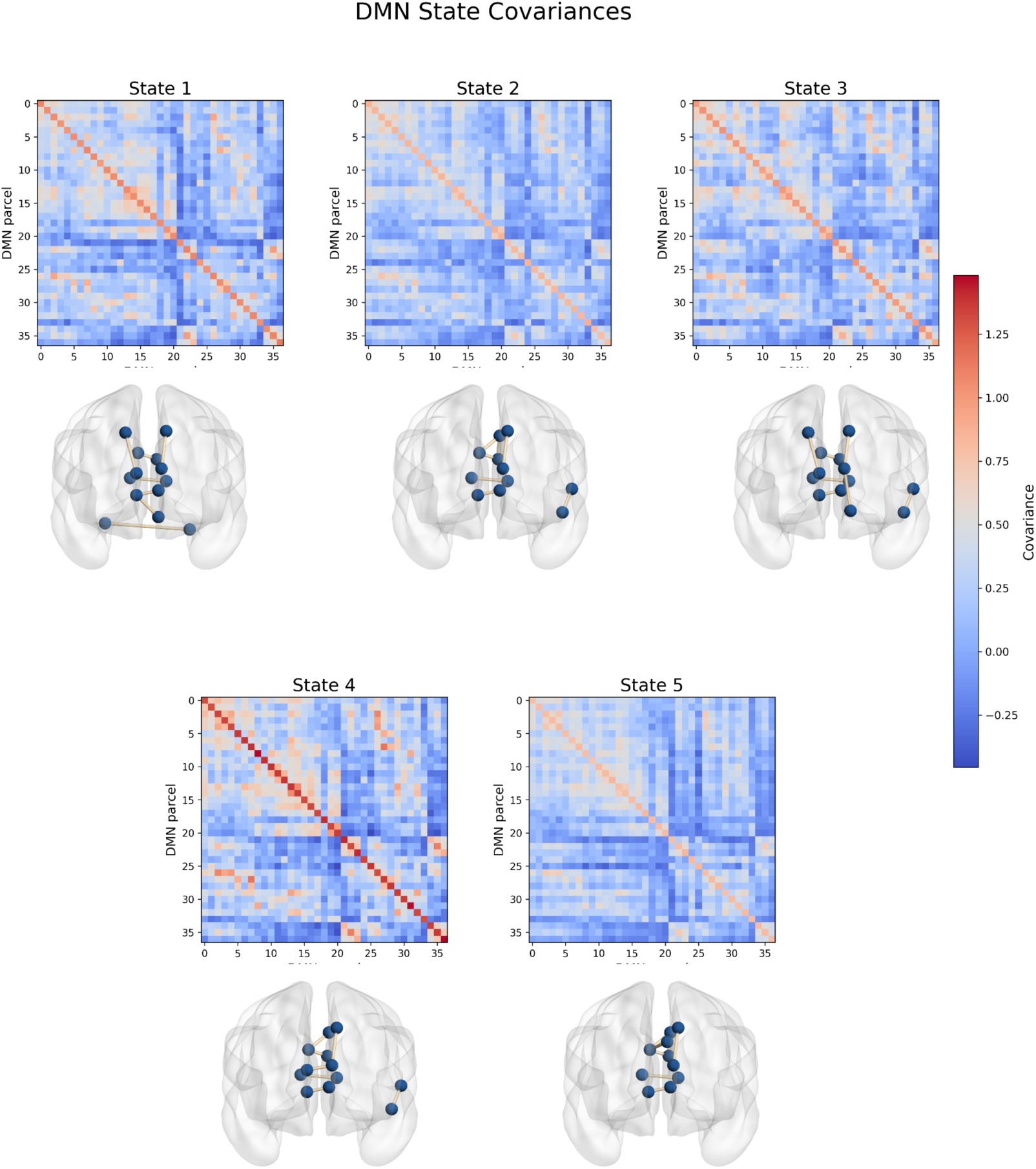
Covariances differ across the five DMN substates. In addition to estimating the mean of each DMN substate (Figure 4), fitting the HMM also yields an estimate of their covariances. For each state, the top panel shows the covariance matrix, with warm colors indicating greater covariance and cooler colors indicating less covariance. The bottom panel shows the pairs of DMN regions in the top 1% of covariance, visualized using BrainNet Viewer (Xia et al., 2013). Compared to the other states, state 4 displayed the highest covariance and state 5 the lowest. Across states, the strongest covariance was generally exhibited by interhemispheric connections between midline DMN regions.

**Supplementary Figure 4.**
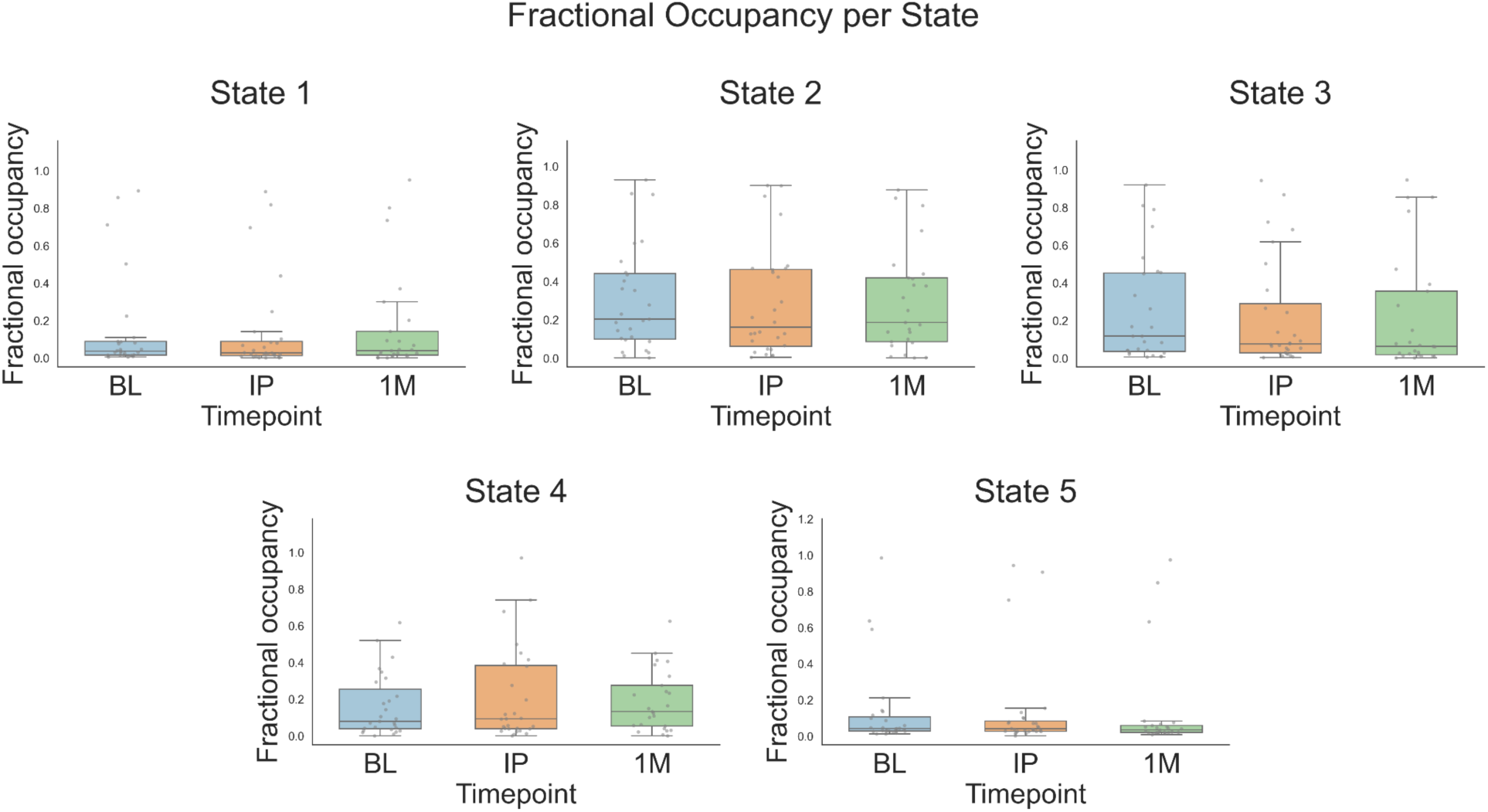
Ibogaine does not have a significant effect on the fractional occupancy of DMN substates. Fractional occupancy is defined as the proportion of total time that the brain spends in a given state. Statistically significant changes in fractional occupancy were determined based on a linear mixed-effects model with timepoint as fixed effect, and then FDR correction was used to adjust for multiple comparisons across the five states and two timepoints. No states exhibited significant changes in fractional occupancy across any of the timepoints.

**Supplementary Figure 5.**
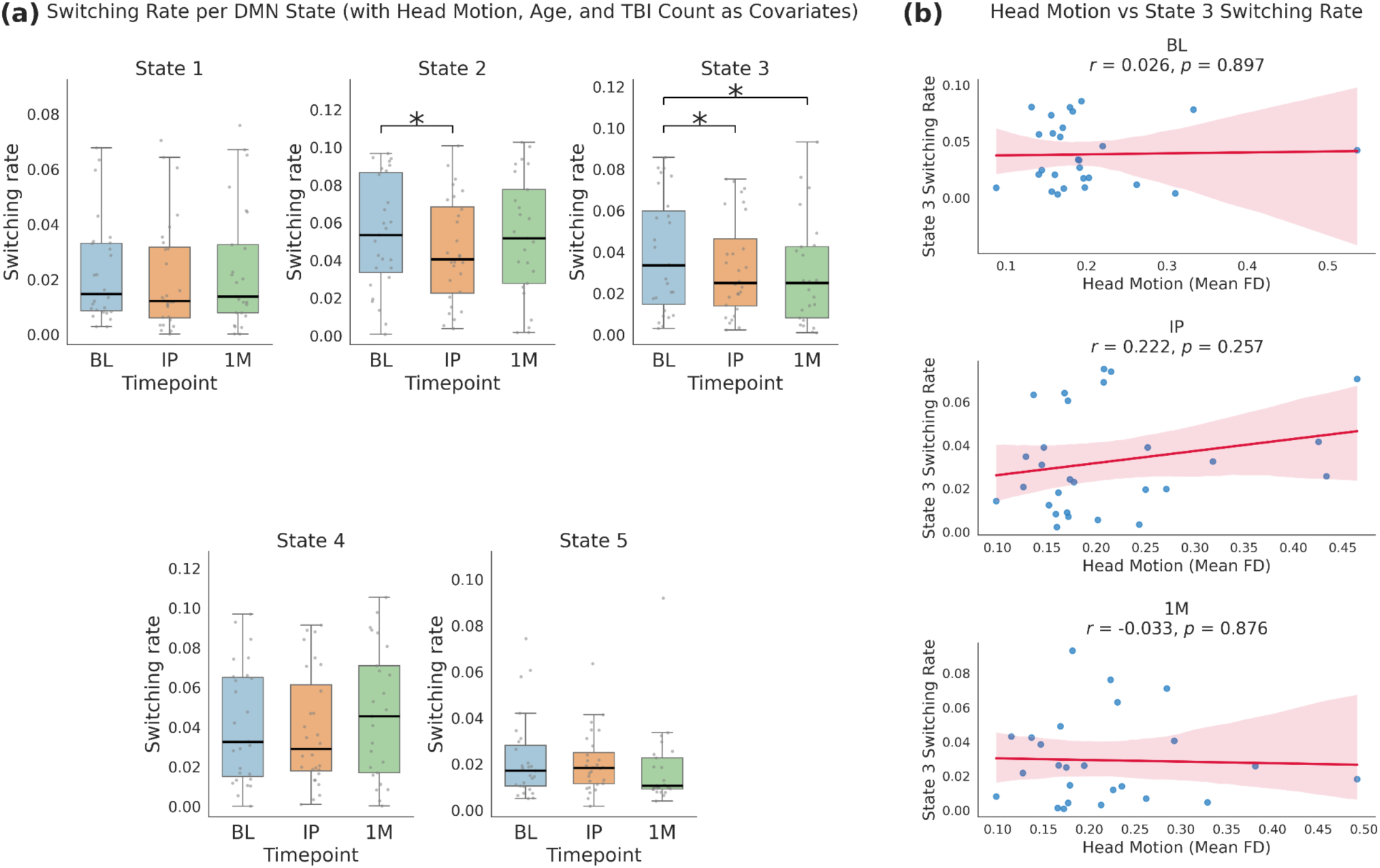
After adjusting for head motion, age, and number of TBIs, ibogaine is still associated with a significant decrease in the switching rate of DMN substate 3 at both timepoints. (a) Head motion was measured with mean framewise displacement (FD) per session, which, together with age and number of TBIs (determined at screening), was included as a fixed effect in the linear mixed-effects model relating timepoint to the switching rate of each state. After multiple comparisons, the decrease in the switching rate of DMN substate 3 was still significant at both IP and 1M. (b) Mean FD was not significantly correlated with the switching rate of DMN substate 3 at any of the three timepoints.

**Supplementary Figure 6.**
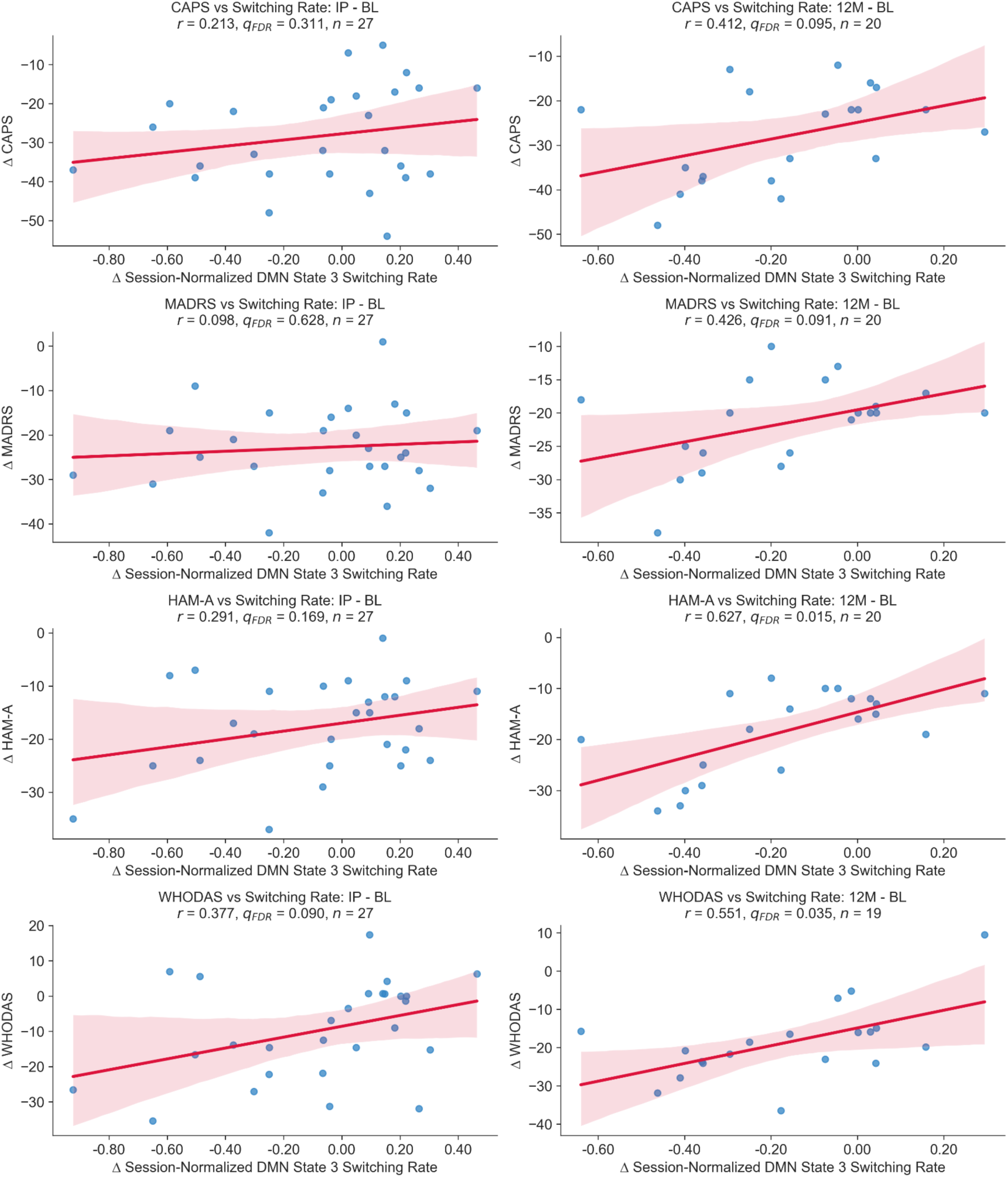
Session-normalized DMN substate 3 switching rate at one month-post ibogaine is positively correlated with some long-term clinical outcomes. To account for inter-individual differences in overall HMM switching propensity, the switching rate of DMN substate 3 per subject per session was divided by the mean switching rate of all five DMN substates for that subject and session. Pearson correlations were computed between session-normalized switching rate of DMN substate 3 at the 1M timepoint and total scores on various clinical scales at IP and 12 months-post ibogaine (12M). Clinical scales included the CAPS-5, which measures PTSD; MADRS, which measures depression; HAM-A, which measures anxiety; and WHODAS, which measures functional disability. After multiple comparisons, HAM-A and WHODAS scores at 12M were significantly and positively correlated with session-normalized switching rate of DMN substate 3 at 1M.

**Supplementary Figure 7.**
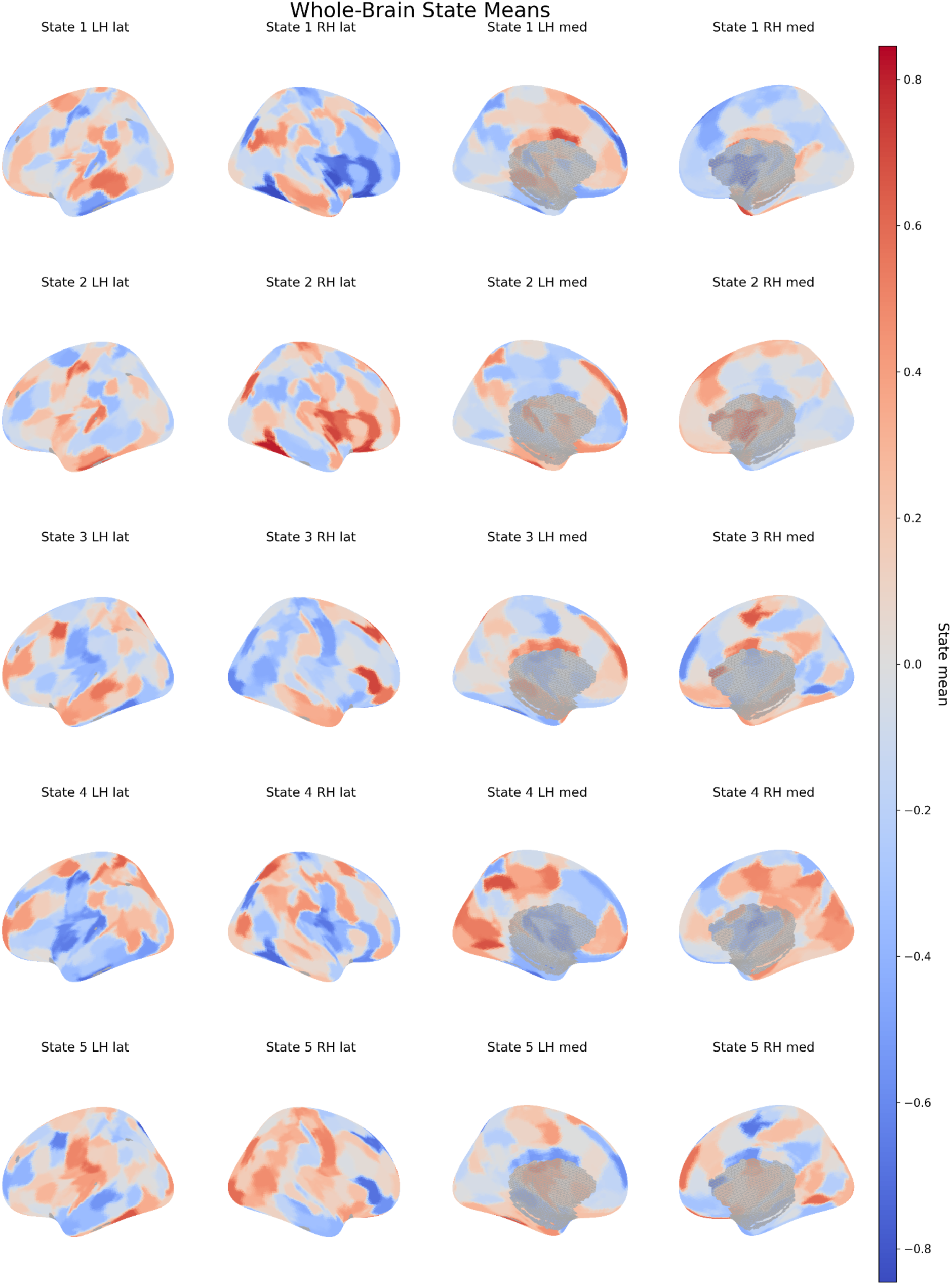
HMM on whole-brain activity reveals large-scale networks. HMMs were also fitted to the fMRI timeseries of the entire brain rather than just the DMN parcels. PCA was used to reduce fMRI data to 15 components and covariance was shared across states. *K* = 5 was found to be the optimal number of states, based on free energy. State means are shown on lateral and medial cortical surfaces for both hemispheres after back-projection from PCA space to parcel space. Warmer colors indicate parcels with relatively higher mean activity for a given state, whereas cooler colors indicate relatively lower mean activity. Among these five states, whole-brain state 3 showed the strongest positive correspondence with the DMN, as defined by the Schaefer parcellation (*r* = 0.25, *p* = 0.0003); whole-brain state 5 showed the strongest negative correspondence (*r* = −0.24, *p* = 0.0004).

**Supplementary Figure 8.**
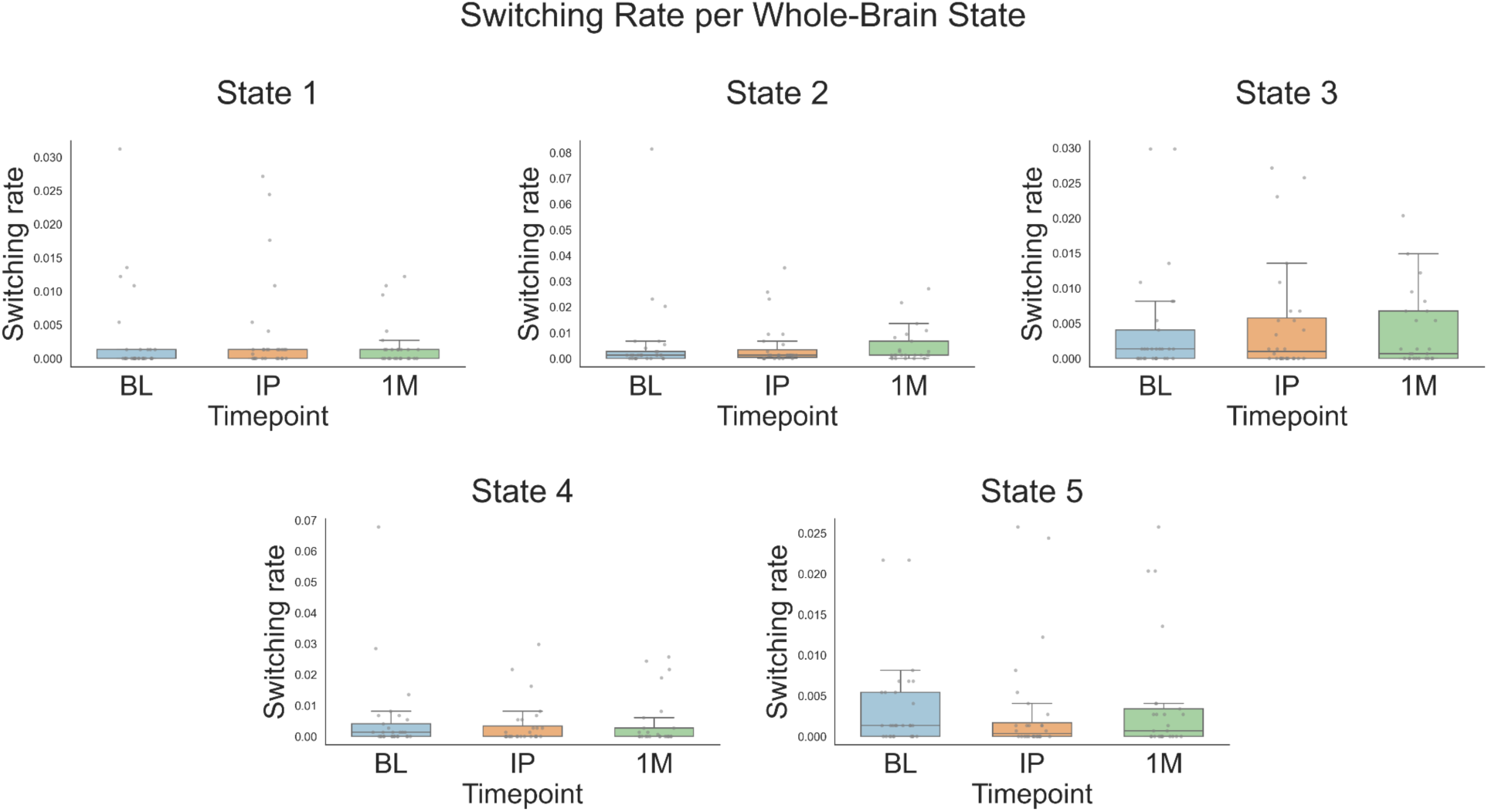
Ibogaine does not have a significant effect on switching rates of whole-brain states. Switching rate is defined as the frequency at which the brain moves from one hidden state to another over time, and it is inversely related to the duration of a state. Statistically significant changes in switching rate of whole-brain HMM states were determined based on a linear mixed-effects model with timepoint as fixed effect, and then FDR correction was used to adjust for multiple comparisons across the five states and two timepoints (IP and 1M). Across states and timepoints, there was no significant effect of ibogaine on switching rate of whole-brain states.

**Supplementary Figure 9.**
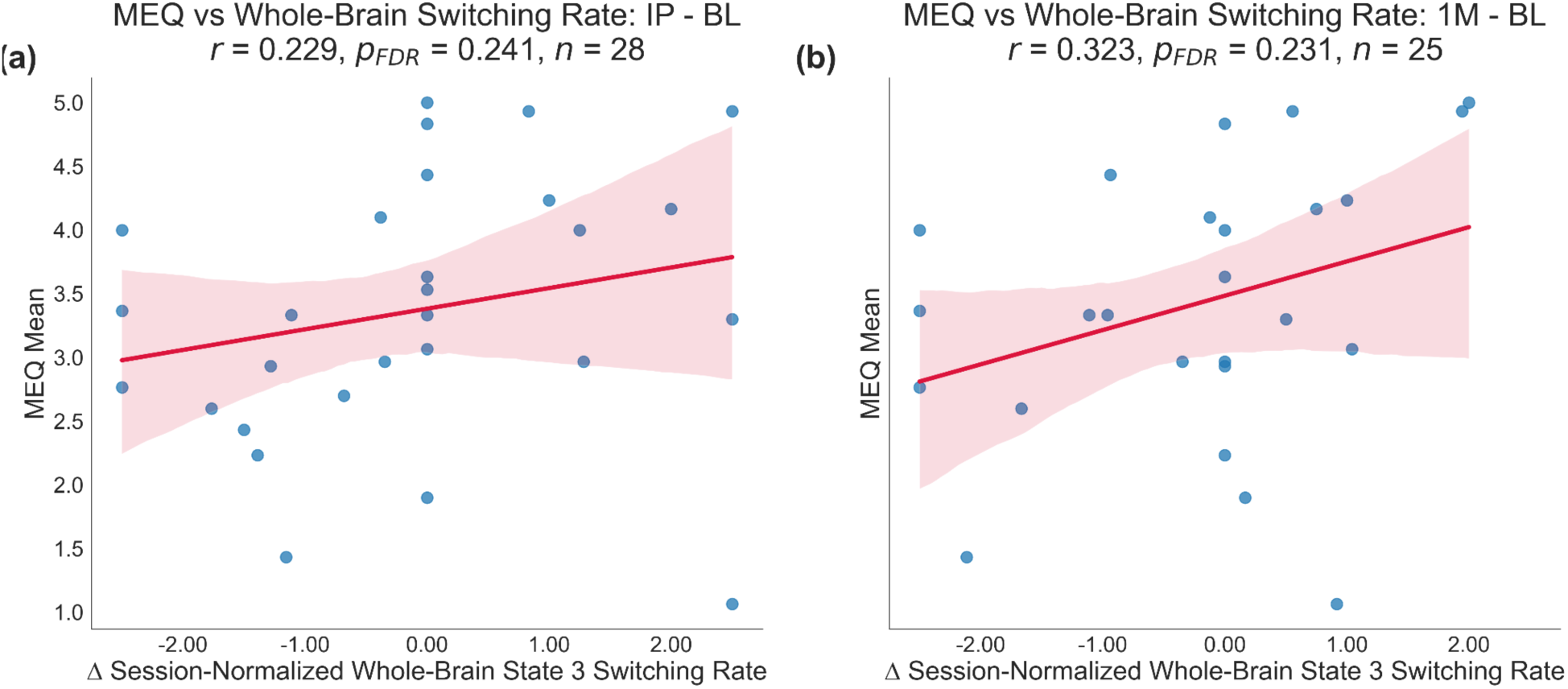
Session-normalized switching rate of whole-brain state 3 is not correlated with the intensity of the mystical experience at either timepoint. Whole-brain state 3 is the state whose mean activation most strongly resembles the DMN. To account for inter-individual differences in overall HMM switching propensity, the switching rate of whole-brain state 3 per subject per session was divided by the mean switching rate of all five whole-brain states for that subject and session. Pearson correlations were conducted between the mean score on the MEQ and the change in the session-normalized switching rate of whole-brain state 3 at both timepoints (IP and 1M). FDR correction was used to adjust for multiple comparisons across the two timepoints. Red shaded bands represent the 95% confidence interval for the estimated mean regression line. The change in whole-brain state 3 switching rate at both timepoints is positively but not significantly correlated with the mean MEQ score.

**Supplementary Figure 10.**
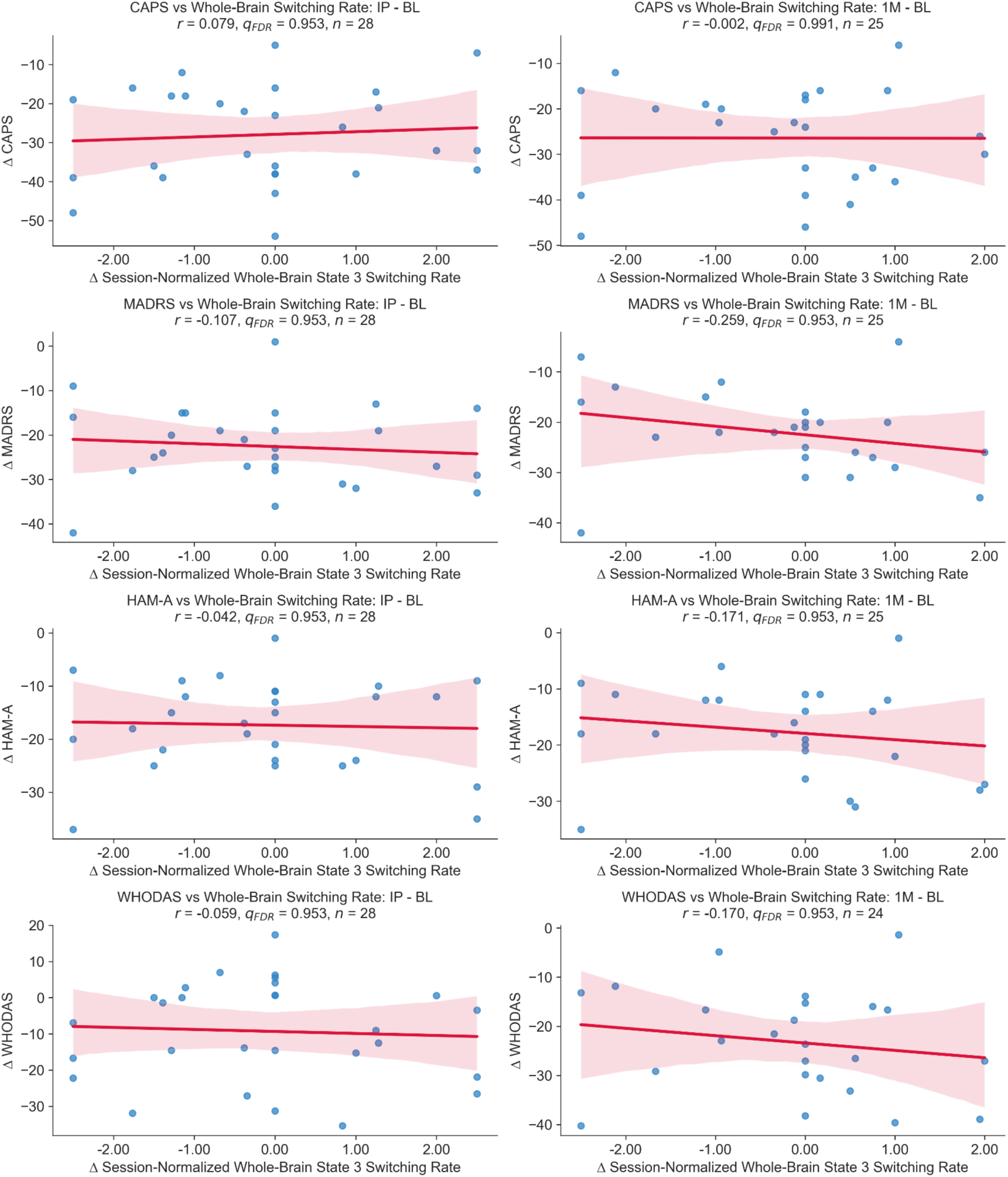
Session-normalized switching rate of whole-brain state 3 is not correlated with clinical improvements at either timepoint. Whole-brain state 3 is the state whose mean activation most strongly resembles the DMN. To account for inter-individual differences in overall HMM switching propensity, the switching rate of whole-brain state 3 per subject per session was divided by the mean switching rate of all five whole-brain states for that subject and session. Pearson correlations were computed between total scores on various clinical scales and session-normalized switching rate of whole-brain state 3. These scales included the CAPS-5, which measures PTSD; MADRS, which measures depression; HAM-A, which measures anxiety; and WHODAS, which measures functional disability. Session-normalized switching rate of whole-brain state 3 was not correlated with any of the clinical outcomes at either timepoint.

**Supplementary Table 1.** Effect of ibogaine on within- and between-network functional connectivity. Statistically significant changes in within-network FC were determined based on a linear mixed-effects model with timepoint as fixed effect, and then false discovery rate (FDR) correction was used to adjust for multiple comparisons across the seven networks. The only change that survived multiple comparisons (bold) was a decrease in the within-network FC of the salience network (SN) at IP. Ibogaine did not have a significant effect on between-network functional connectivity for any pairs of networks.

| Analysis | Network(s) | Contrast | Beta | SE | p | q (BH-FDR) |
| --- | --- | --- | --- | --- | --- | --- |
| Within-network | Cont | 1M vs BL | -0.001 | 0.010 | 0.954 | 0.954 |
| Within-network | Cont | IP vs BL | -0.004 | 0.009 | 0.705 | 0.882 |
| Within-network | Default | 1M vs BL | -0.028 | 0.012 | 0.025 | 0.175 |
| Within-network | Default | IP vs BL | -0.016 | 0.012 | 0.168 | 0.438 |
| Within-network | DorsAttn | 1M vs BL | -0.022 | 0.017 | 0.188 | 0.438 |
| Within-network | DorsAttn | IP vs BL | -0.024 | 0.016 | 0.136 | 0.438 |
| Within-network | Limbic | 1M vs BL | -0.005 | 0.017 | 0.756 | 0.882 |
| Within-network | Limbic | IP vs BL | -0.034 | 0.017 | 0.041 | 0.191 |
| Within-network | SN | 1M vs BL | -0.024 | 0.020 | 0.232 | 0.464 |
| Within-network | SN | IP vs BL | -0.064 | 0.020 | 0.001 | **0.016** |
| Within-network | SomMot | 1M vs BL | -0.032 | 0.030 | 0.277 | 0.485 |
| Within-network | SomMot | IP vs BL | -0.017 | 0.029 | 0.541 | 0.757 |
| Within-network | Vis | 1M vs BL | 0.004 | 0.020 | 0.840 | 0.905 |
| Within-network | Vis | IP vs BL | 0.013 | 0.019 | 0.502 | 0.757 |
| Between-network | Cont - Default | 1M vs BL | -0.179 | 0.118 | 0.129 | 0.531 |
| Between-network | Cont - Default | IP vs BL | 0.132 | 0.114 | 0.248 | 0.531 |
| Between-network | DorsAttn - Cont | 1M vs BL | -0.209 | 0.155 | 0.178 | 0.531 |
| Between-network | DorsAttn - Cont | IP vs BL | 0.071 | 0.150 | 0.635 | 0.740 |
| Between-network | DorsAttn - Default | 1M vs BL | -0.206 | 0.128 | 0.108 | 0.531 |
| Between-network | DorsAttn - Default | IP vs BL | 0.077 | 0.125 | 0.538 | 0.718 |
| Between-network | DorsAttn - Limbic | 1M vs BL | -0.346 | 0.142 | 0.015 | 0.316 |
| Between-network | DorsAttn - Limbic | IP vs BL | -0.147 | 0.138 | 0.286 | 0.531 |
| Between-network | DorsAttn - SN | 1M vs BL | -0.203 | 0.166 | 0.223 | 0.531 |
| Between-network | DorsAttn - SN | IP vs BL | 0.044 | 0.161 | 0.783 | 0.843 |
| Between-network | Limbic - Cont | 1M vs BL | -0.313 | 0.143 | 0.029 | 0.405 |
| Between-network | Limbic - Cont | IP vs BL | -0.055 | 0.139 | 0.692 | 0.785 |
| Between-network | Limbic - Default | 1M vs BL | -0.305 | 0.126 | 0.015 | 0.316 |
| Between-network | Limbic - Default | IP vs BL | -0.068 | 0.121 | 0.574 | 0.718 |
| Between-network | SN - Cont | 1M vs BL | -0.159 | 0.154 | 0.304 | 0.531 |
| Between-network | SN - Cont | IP vs BL | 0.105 | 0.150 | 0.483 | 0.698 |
| Between-network | SN - Default | 1M vs BL | -0.149 | 0.130 | 0.253 | 0.531 |
| Between-network | SN - Default | IP vs BL | 0.126 | 0.126 | 0.320 | 0.538 |
| Between-network | SN - Limbic | 1M vs BL | -0.253 | 0.167 | 0.129 | 0.531 |
| Between-network | SN - Limbic | IP vs BL | -0.014 | 0.163 | 0.929 | 0.961 |
| Between-network | SomMot - Cont | 1M vs BL | -0.191 | 0.175 | 0.274 | 0.531 |
| Between-network | SomMot - Cont | IP vs BL | 0.062 | 0.169 | 0.713 | 0.788 |
| Between-network | SomMot - Default | 1M vs BL | -0.176 | 0.138 | 0.200 | 0.531 |
| Between-network | SomMot - Default | IP vs BL | 0.074 | 0.133 | 0.581 | 0.718 |
| Between-network | SomMot - DorsAttn | 1M vs BL | -0.267 | 0.190 | 0.161 | 0.531 |
| Between-network | SomMot - DorsAttn | IP vs BL | -0.014 | 0.185 | 0.938 | 0.961 |
| Between-network | SomMot - Limbic | 1M vs BL | -0.312 | 0.172 | 0.070 | 0.531 |
| Between-network | SomMot - Limbic | IP vs BL | -0.083 | 0.168 | 0.620 | 0.740 |
| Between-network | SomMot - SN | 1M vs BL | -0.194 | 0.174 | 0.267 | 0.531 |
| Between-network | SomMot - SN | IP vs BL | 0.002 | 0.168 | 0.993 | 0.993 |
| Between-network | Vis - Cont | 1M vs BL | -0.125 | 0.121 | 0.301 | 0.531 |
| Between-network | Vis - Cont | IP vs BL | 0.162 | 0.117 | 0.166 | 0.531 |
| Between-network | Vis - Default | 1M vs BL | -0.137 | 0.101 | 0.175 | 0.531 |
| Between-network | Vis - Default | IP vs BL | 0.150 | 0.098 | 0.125 | 0.531 |
| Between-network | Vis - DorsAttn | 1M vs BL | -0.121 | 0.128 | 0.346 | 0.559 |
| Between-network | Vis - DorsAttn | IP vs BL | 0.084 | 0.124 | 0.498 | 0.698 |
| Between-network | Vis - Limbic | 1M vs BL | -0.191 | 0.119 | 0.107 | 0.531 |
| Between-network | Vis - Limbic | IP vs BL | -0.064 | 0.115 | 0.579 | 0.718 |
| Between-network | Vis - SN | 1M vs BL | -0.098 | 0.133 | 0.462 | 0.693 |
| Between-network | Vis - SN | IP vs BL | 0.160 | 0.129 | 0.216 | 0.531 |
| Between-network | Vis - SomMot | 1M vs BL | -0.105 | 0.139 | 0.448 | 0.693 |
| Between-network | Vis - SomMot | IP vs BL | 0.141 | 0.135 | 0.296 | 0.531 |

